# Wastewater Genomic Surveillance Captures SARS-CoV-2 Early Detection, Cryptic Transmission, and Variant Dynamics

**DOI:** 10.1101/2025.11.27.25332381

**Authors:** G. Veytsel, A. Sullivan, L. Lariscy, M. Lott, E. Lipp, T. Glenn, L. Carmola, H. Dishman, J. Bahl

## Abstract

Wastewater-based epidemiology addresses key biases and limitations in clinical surveillance by providing population-level coverage, capturing subclinical infections, and mitigating limited clinical testing. Despite its demonstrated value during the COVID-19 pandemic, few phylogenetic studies of wastewater sequences have been conducted, with inferences limited to descriptive genomic diversity. Phylogenetic methods offer deeper insights into viral ancestry, transmission, and epidemic dynamics, yet their application to wastewater data remains unexplored. Here, we present the first study to apply Bayesian phylodynamic approaches to characterize SARS-CoV-2 variant dynamics using wastewater sequences.

We conducted a 2.5-year long longitudinal study (June 2020 – December 2022) using wastewater samples collected from catchment sites in Clarke County, Georgia. Leveraging an optimized enrichment and sequencing strategy, we generated 80 whole genome sequences from 41 wastewater samples. As proof of concept for combining wastewater and clinical sequences, we reconstructed epidemic waves in Clarke County in the context of national and international transmission.

Phylogenetic analyses showed that wastewater sequences captured all clinically observed variants, inferred variant emergence times consistent with or earlier than clinical genomes, and revealed cryptic Beta transmission. We further identified associations between student movement events and clade expansion, increased transmission, and subsequent case surges. Together, these results demonstrate the power of wastewater genomics to reconstruct viral emergence and spread. Integrating wastewater and clinical sequences provides a robust phylogenetic framework for identifying cryptic transmission, inferring epidemic history, and contextualizing local viral circulation within broader transmission patterns. These findings underscore the importance of wastewater surveillance for pandemic preparedness and public health response.

## Introduction

The evolution and emergence of severe acute respiratory syndrome coronavirus 2 (SARS-CoV-2) variants, such as Alpha, Delta, and Omicron, have caused global surges during the coronavirus disease 2019 (COVID-19) pandemic (1). These variants have accumulated a defining set of mutations that can affect transmissibility, disease severity, diagnostics, and the effectiveness of treatments and vaccines (2). Therefore, timely identification of SARS-CoV-2 variants is critical for protecting communities. Infected individuals can shed SARS-CoV-2 in feces, urine, sputum, saliva, and blood, all of which may enter the sewer system at different magnitudes and probabilities. Feces and sputum, however, contribute most to SARS-CoV-2 RNA concentrations detected in wastewater (3,4). While wastewater-based epidemiology (WBE) emerged as a key tool during the COVID-19 pandemic, it has been employed for community monitoring of many viral pathogens for decades, including poliovirus (5), influenza virus (6), respiratory syncytial virus (7), enterovirus (8), and mpox virus (9,10).

The key advantages of wastewater for population-wide surveillance lie in its widespread coverage: in the United States, 80% of households are served by municipal sewage systems (11). Therefore, wastewater surveillance captures asymptomatic and pre-symptomatic cases, which are likely missed by clinical surveillance. Consequently, WBE represents a more comprehensive snapshot of COVID-19 burden in communities, which is important in refining estimates such as case-fatality rates, informing models of virus transmission dynamics, evaluating interventions, and aiding public health decision-making (e.g., when to lift restrictions). WBE can further serve as an early warning system for emerging variants and can identify rapidly worsening areas to provide valuable time for targeted public health interventions. In many cities, SARS-CoV-2 RNA was detected in wastewater weeks before the first confirmed COVID-19 case (12–14). Because wastewater surveillance is independent of healthcare-seeking behaviors and availability/accessibility of clinical testing, it minimizes biases associated with clinical surveillance. Wastewater surveillance is a low-cost (15), time-efficient, and non-invasive strategy that can provide continuous community surveillance, in addition to focused monitoring of high-risk populations or areas with a high infection burden (11).

During the first few years of the COVID-19 pandemic, wastewater surveillance focused on detecting and quantifying SARS-CoV-2 viral loads using PCR-based approaches. As sequencing techniques improved, researchers began recovering complete and nearly complete SARS-CoV-2 genomes from wastewater (16,17). Unlike clinical sequences that are comprised of a single variant, wastewater samples are comprised of multiple lineages or variants. Therefore, computational tools for deconvolution such as Freyja (18), Cojac (19), and Kallisto (20) were developed to assign reads to SARS-CoV-2 lineages and estimate the relative abundance of different lineages in mixed samples. These methods allowed researchers to monitor variant dynamics. Early genomic studies were largely limited to describing SARS-CoV-2 comparative genetic diversity. Studies showed that variant abundance and dynamics observed in wastewater closely correlated with variant profiles from clinical sequencing (16,17,21,22). Wastewater surveillance also identified cryptic transmission (i.e., variants not detected by clinical genomic surveillance) and identified emerging variants up to two weeks before being captured by clinical surveillance(18,19).

There are few phylogenetic studies of wastewater-derived sequences. These studies have very small sample sizes (one to 22 sequences) and are limited to tree-building for further comparative genetic diversity characterization of wastewater and clinical SARS-CoV-2 genomes (17,23,24). However, phylogenetic approaches can be a powerful tool to reconstruct viral ancestry, track the spread of SARS-CoV-2 variants, examine source-sink dynamics, and infer epidemiological patterns, such as the reproductive number, viral evolutionary rates, or virus effective population sizes. Still, the effectiveness of using wastewater sequences to infer epidemic dynamics and transmission history should be evaluated. To our knowledge, this is the first study that uses a Bayesian phylodynamic approach to study sequences obtained from wastewater.

In this study, we conducted a longitudinal investigation, collecting wastewater samples across 2.5 years (June 2020 – December 2022) from three catchment sites in Clarke County, Georgia. Clarke County is home to Athens, Georgia, a college town with population changes driven by the seasonal influx of ∼30,000 students. This migration pattern offers a unique opportunity to document the arrival and exportation of SARS-CoV-2 variants from around the state and nation with student movement events. We began with a comparative phylogenetic analysis using wastewater and clinical sequences to explore if wastewater sequences corroborate the results and inferences found in clinical surveillance. We explored the unique inferences provided by wastewater data and interrogated the limitations of wastewater sequences in reconstructing epidemic dynamics. For example, we assessed if wastewater sequences reveal novel variants or unobserved dynamics compared to clinical sequences. We also examined if combining wastewater and clinical data provides a more comprehensive picture of variant dynamics. Lastly, we applied spatial diffusion models to a dataset of clinical and wastewater genomes to understand transmission dynamics within the context of important student movement dates.

## Materials and Methods

### Wastewater sampling

For wastewater surveillance of SARS-CoV-2, we sampled across three sewersheds in Athens-Clarke County, GA, between June 2020 – December 2022, representing a total population of about 130,000 people. We evaluated about 700 composite samples for SARS-CoV-2, representative of the average viral signal over the 24-hour period prior to sample retrieval (25,26).

### RNA extraction for wastewater samples

Solid pellets (∼ 1 mL) were concentrated from each wastewater sample (100 mL) and prepared for RNA purification. Samples were purified using the Zymo Environ Water RNA Kit following manufacturer instructions, with minor modifications. Following sample homogenization via bead beating, RNA was extracted from the supernatant and filtered.

### Library preparation of wastewater samples

The most accurate and effective library preparation method for short read sequencing of SARS-CoV-2 derived from wastewater was determined in a previous study (27). The protocol is available on protocols.io (https://dx.doi.org/10.17504/protocols.io.rm7vzj6zrlx1/v1). Initial targeted amplification was performed using the commercially available Integrated DNA Technologies (IDT) ARTIC V4.1 tiled amplicon panel, followed by amplification with IDT custom-ordered fusion primers. These primers follow the same amplicon scheme and are designed to facilitate tagmentation of each library. This approach enables further amplification of target molecules while eliminating the need for end repair and adapter ligation. Completion of this protocol should result in a clean, multiplexed DNA library pool ready for sequencing. A key feature of this protocol is the addition of custom fusion primers, which allow for large scale-multiplexing of barcodes, along with multiple cleaning steps.

### Sequencing of wastewater samples

320 wastewater samples were sequenced using previously identified SARS-CoV-2 specific library preparation methods (26). Libraries were pooled to achieve 500,000 paired reads per sample and sequenced on an Illumina NovaSeq 6000 with PE250 reads. All wastewater samples were pooled in a single run to avoid variation between sequencing runs. The entire genome was sequenced with 100x coverage to ensure accurate variant assignment. Base calling accuracy was measured using Phred quality scores; Phred scores of 20 correspond to 99% precision (28). Adapters and bases that had a Phred Score < 20 were trimmed from the reads and reads of length 100 base pairs (bp) or less were removed in Trimmomatic (29). 129 wastewater samples obtained sufficient reads for further analyses.

### Genome assembly of wastewater reads

Given that each sequenced sample contains a mixture of SARS-CoV-2 strains circulating within wastewater, we used the StrainSort pipeline (https://github.com/mandysulli/StrainSort_pipeline) to deconvolute lineages and perform de novo assembly (30). Kallisto (20) was used to assign reads to a strain and estimate lineage abundance. All sequenced samples contained sufficient reads to estimate variant abundance of different strains within the sample. Reads were then separated by strain using the separation function in StrainSort and assigned to strain-specific FASTQ files. Reads that did not align to any known strain but were identified as “SARS-CoV-2” were retained in a separate FASTQ file and included in downstream analyses. This approach facilitated the identification of emerging strains within wastewater samples. De novo assembly of reads by strain was performed in SPAdes (31). Java programs were developed in StrainSort to perform the following tasks: filtering assembled genomes to exclude genomes with a high proportion of Ns (>50%) in the spike protein region and constructing a consensus genome for each strain using scaffolds generated by SPAdes. This process ensured that all genomes had a uniform length and were aligned to each other, resulting in 80 whole genome sequences from 41 samples. These sequences were then used for multiple sequence alignment for downstream phylogenetic analyses.

### Clinical sequences

Whole genome sequences of SARS-CoV-2 obtained from human COVID-19 cases in Georgia were downloaded from the Global Initiative on Sharing Avian Influenza Data-EpiCoV (GISAID-EpiCoV) platform (https://www.gisaid.org/) (32). Only sequences submitted up to December 7, 2022 with complete genomes (greater than 29,000 nucleotides) and complete collection dates were included. Linked zip code metadata were provided by the Georgia Department of Public Health (GA DPH). The zipcodeR package in R (34) was used to assign zip codes to county. Clinical sequences were filtered to Clarke County during the same time period as the wastewater samples (2021-04-14 to 2022-09-05), resulting in 632 sequences.

To generate a contextual dataset for the spatial analyses, a random sample of 100,000 genomes from all global sequences as of November 16, 2024 was downloaded from GISAID. Only sequences collected during the same time period as the wastewater samples (2021-04-14 to 2022-09-05) were queried. Previous criteria were applied: human host, complete genome, and complete collection date. Georgia contextual sequences were linked to zip codes from GA DPH and assigned to county and public health district. Public health districts are an administrative boundary for health policy and funding, based on the following designations: https://dph.georgia.gov/public-health-districts. Georgia contextual sequences were subsampled proportionately by collection year, month, and public health district in order to generate a spatially and temporally representative subsample (n = 182). In addition, global contextual sequences were subsampled proportionately by collection year, month, and region (n = 108).

All PCR confirmed and antigen-positive cases, by county and week, were downloaded from the Georgia Department of Health (https://dph.georgia.gov/covid-19-status-report) as of January 16, 2025.

### Alignment of wastewater and clinical samples

Reference based alignment of clinical sequences was generated in MAFFT (33) against the Wuhan reference (GenBank accession MN908947). While some of our wastewater sequences were low coverage, they contained biologically important information. To mitigate negative effects from potential noise on the quality of our wastewater multiple sequence alignment (MSA), we selected highly reliable sequences (Clarke County clinical sequences) to build a backbone MSA. Wastewater sequences were added to the backbone alignment thereby conserving positional homology and reducing alignment error. As a result, the quality of the final MSA was less affected by potential quality issues in the wastewater sequences (33). We kept these clinical sequences in our wastewater MSA for our combined phylogeny to compare parameter estimates and distribution against wastewater and clinical data alone.

### Alignment optimization

De novo assembly requires aggressive error correction due to the fragmented and partial nature of the genomes being assembled. To improve alignment accuracy, an algorithm was applied in R to identify and remove regions likely representing misassembled sequences (chimers). Since SARS-CoV-2 sequences have high sequence similarity, large deviations are not expected. Consecutive base changes are rare and indels are usually very small (<3 nucleotides) (34–36); therefore, a conservative threshold of 40% genomic dissimilarity was set to detect potential misassemblies. This algorithm was based on the Levenshtein distance to measure the number of differences (i.e., insertions, deletions, and substitutions) between each wastewater sequence and the Wuhan reference genome. If the distance was greater or equal to four in each 10-base sliding window, the corresponding portion of the wastewater sequence was replaced with gaps. All MSAs were manually inspected and trimmed to include only coding regions.

### Phylogenetic analyses

ModelFinder as implemented in the IQ-TREE software (37), determined the best fit model to be the general time reversible (GTR) model with unequal base frequencies empirically computed from the MSA (F), with invariant sites allowed (I), and four categories of site-rate heterogeneity under the Gamma model (G4). Based on the maximum likelihood tree produced by IQ-TREE v2.2 (40), a root-to-tip regression was performed using TempEst v1.5.3 (38) as a preliminary assessment of the presence of temporal signal within the sequence data (Supplementary Figure 1). Bayesian phylogenetic trees were estimated using BEAST v1.10.4 (39) with an uncorrelated relaxed lognormal clock model that allows for evolutionary rate variation across branches. The skyride demographic model was specified to investigate population dynamics. Per the early estimated rate of evolution (40), the prior mean rate for the relaxed, lognormal molecular clock model was set at 8 x 10^-4^ substitutions/site/year, under a lognormal distribution with a standard deviation of 1 x 10^-3^. At least three independent MCMC runs of 200 million chain length were performed from random starting trees. Convergence and chain stationarity were assessed in Tracer (41), ensuring an ESS > 200 for each estimated parameter after removing burn-in. Tree and log files were combined and resampled in LogCombiner (39) to achieve 10,000 samples. This workflow was used for analyses for Clarke County clinical sequences, Clarke County wastewater sequences, and Clarke County combined datasets (wastewater and clinical). A maximum clade credibility (MCC) tree was obtained using TreeAnnotator v1.10.4 (39) for each cluster and visualized in R.

To characterize introductions and source-sink dynamics between Clarke County and the rest of the state, we combined all wastewater sequences from Clarke County (n=80) with a subsample of clinical Clarke County sequences (n=261). Clarke County sequences were contextualized with worldwide sequences in a 1:1 ratio. Contextual data were comprised of a subsample from Georgia (n=182) and worldwide sequences (n=109), proportional to the number of sequences in each public health district and region, respectively. All subsamples were also temporally representative, grouped by year and month of collection. Discrete trait analysis was conducted using these empirical set of trees based on three traits: Clarke County, Georgia, and out of state. A Bayesian Stochastic Search Variable Selection (BSSVS) social network was applied to capture location exchange rates that adequately explain the diffusion process (42) and an asymmetric continuous-time Markov chain (CTMC) model inferred migration rates of viral lineages between geographic locations. Bayes Factor (BF) values were calculated using SpreaD3 (43) to assess the support for each pairwise rate of diffusion between locations. To classify the level of support, we used common thresholds in the field (44,45): No support BF < 3; substantial support: BF = 3-10; strong support: BF = 11-30; very strong support: BF = 31-100; decisive support: BF > 100. Five replicates were performed to assess the robustness of our results to resampling.

Despite excellent convergence for relevant parameters, analyses that combined wastewater and clinical sequences consistently had several metrics with an Effective Sample Size (ESS) under 200: standard deviation of the uncorrelated log-normal relaxed clock (ucld.stdev), coefficient of variation, and covariance. In addition to these, treelength for several replicates of the spatial phylogenetic analysis also had ESS values < 200.

### TMRCA estimation

Across all phylogenetic analyses, taxon sets based on shared SARS-CoV-2 variants across wastewater and clinical samples were defined and the time of the most recent common ancestor (TMRCA) was estimated for each variant. These consisted of Alpha (B.1.1.7 and Q lineages), Delta (B.1.617.2 and AY lineages), and Omicron (B.1.1.529 and descendant lineages). By selecting “stem”, BEAST uses each taxon set to specify a subtree (i.e., a clade or a TMRCA). The subtree includes the stem branch above the clade or TMRCA, and is not constrained, so that other taxa can be added into the clade (which is especially relevant for the unmapped wastewater sequences). Priors for each Variant of Concern (VOC) TMRCA were selected based on the literature: Alpha (46), Delta (47), and Omicron (48). While we created a subtree containing all Omicron sublineages in our phylogeny, the resulting MCC tree revealed distinct clades among Omicron descendants. Therefore, we used TreeStat (39) to calculate the posterior distribution of TMRCAs for BA.1, BA.2, BA.4, and BA.5 from our 10,000 trees. The 95% Bayesian Credible Intervals (BCI) of the divergence times were estimated using the HDInterval package in R (49).

## Results

### Comparative genomic analysis of SARS-CoV-2 sequence variants across sewage and clinical sources in Clarke County, GA

A total of 80 wastewater whole genome sequences were recovered from the 41 wastewater samples collected from Clarke County wastewater treatment plants between April 14, 2021 and September 5, 2022. Relative to the Wuhan reference sequence, these sequences had 22.3% to 99.3% coverage and 1-81 mutations, representing a broad range of sequence quality and genomic diversity within the dataset, respectively.

Additionally, we obtained 632 clinical sequences from Clarke County during this time period. These sequences were comprised of high-quality genomes (93.6% to 99.3% coverage) with an accumulated number of mutations over time consistent with the wastewater sequences (0-80 mutations relative to the Wuhan reference). We reconstructed a time-scaled phylogenetic tree of wastewater sequences and clinical sequences to compare inferences. We found that Alpha, Delta, and Omicron were reliably detected across clinical and wastewater sequences. Wastewater sampling identified Beta sequences, which were undetected in clinical surveillance during the study period in Clarke County (Figure 1). This suggests cryptic transmission of Beta during this time period. The wastewater phylogeny also presented unmapped sequences (Figure 1A). These correspond to reads that did not map to anything in the reference database, meaning they could be known strains that were not included or emerging novel variants. Importantly, all unmapped sequences clustered tightly with a known VOC in the tree, so they were likely just excluded known strains.

**Figure 1.**
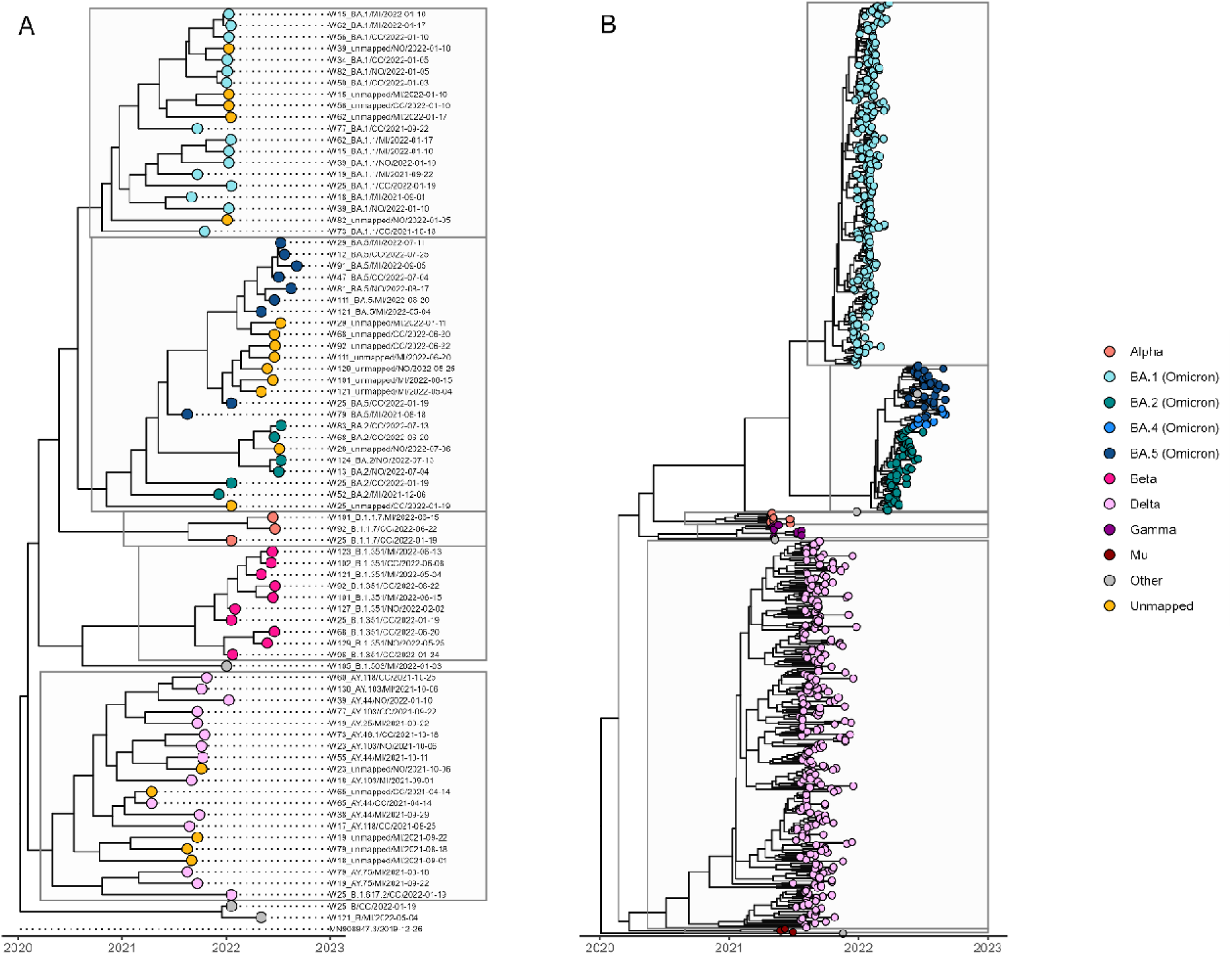
Wastewater sequences identified variants present in clinical samples, plus cryptic Beta transmission. (A) Sequences were collected across three sewersheds between April 27, 2021 and September 3, 2022 (n = 80). Tips are colored by variant, including Alpha, Beta, Delta, and Omicron (BA.1, BA.2, and BA.5). Other includes the Wuhan reference, B.1.503 and B lineages (n=4). Unmapped sequences correspond to reads that did not map to anything in the reference database, meaning they could be known strains that were not included or emerging novel variants. (B) Clinical sequences were collected during the same time period as the wastewater sequences (April 14, 2021 to September 5, 2022) (n = 632). Tips are colored by variant, including Alpha, Delta, Gamma, Mu, and Omicron (BA.1, BA.2, BA.4, and BA.5). Other includes the Wuhan reference, B, B.1.1.409, BE.3, and R.1 lineages (n=6).

### Validation of the use of sequences from wastewater as a population-based surveillance tool to follow infection trends and viral transmission

To examine if combining different sources of data would strengthen inferences from wastewater or clinical sampling alone, we reconstructed a phylogenetic tree from all wastewater and clinical sequences in Clarke County during our study period (n = 712) (Figure 2). In the combined phylogeny, we found that wastewater sequences clustered with clinical sequences within each VOC clade. Sequence diversity from wastewater was representative of clinical cases in the same community; we were able to detect all variants present in the clinical samples, and sufficiently capture the major VOCs Alpha, Delta, and Omicron throughout their period of occurrence in Clarke County.

**Figure 2.**
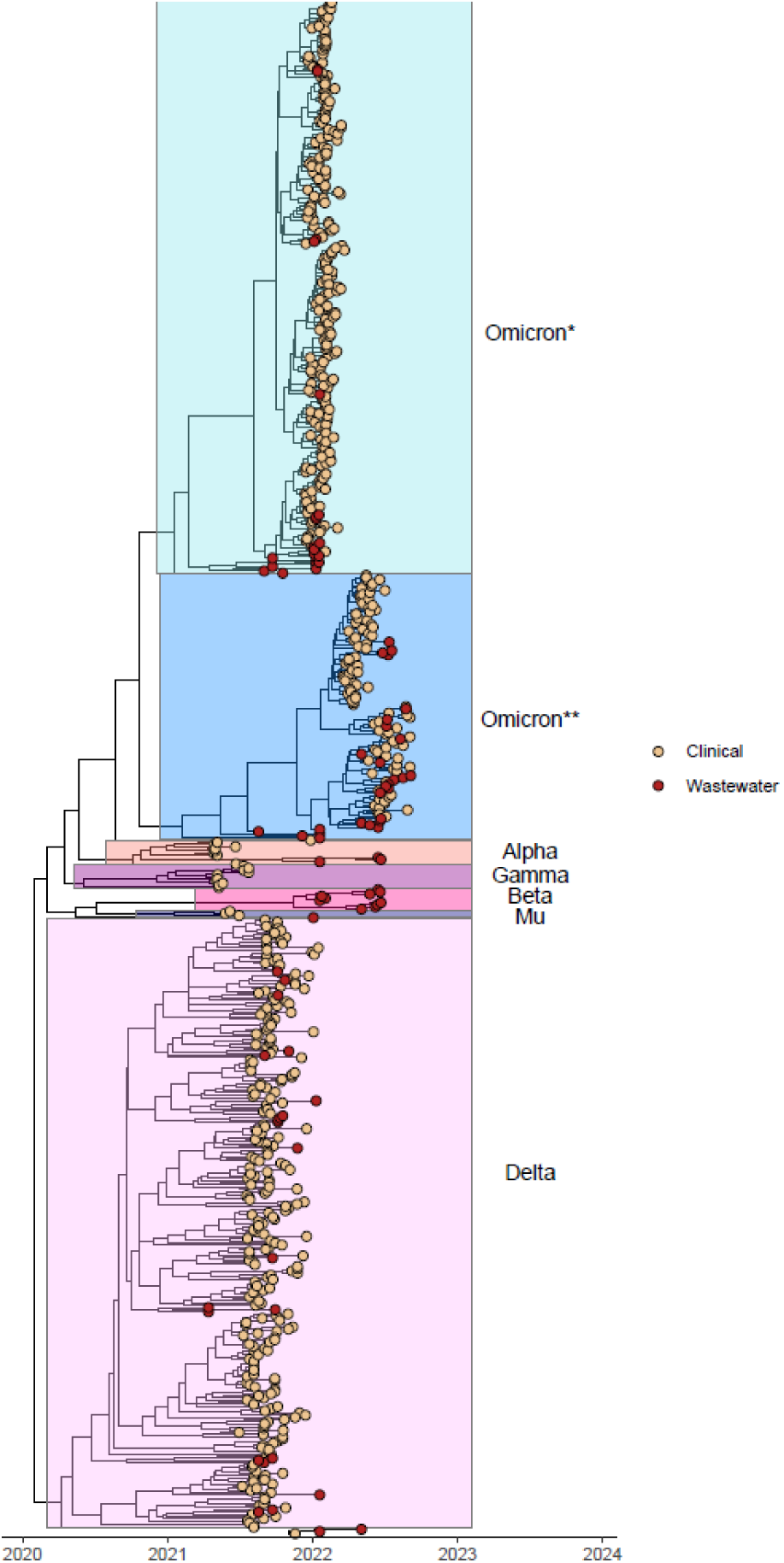
Wastewater sequences clustered with clinical sequences within each VOC clade. Sequences were collected during April 14, 2021 to September 5, 2022 (n=712). Tips are colored by variant, including Alpha, Beta, Delta, and Omicron (BA.1, BA.2, BA.4, and BA.5). Among Omicron sublineages, BA.1 forms a monophyletic clade (Omicron*), while BA2, BA.4 and BA.5 form a paraphyletic clade (Omicron**). Other includes the Wuhan reference, B, B.1.1.409, B.1.503, BE.3, and R.1 lineages (n=8).

In our dataset, we found that the detection of Alpha in wastewater, on January 19, 2022, lagged behind the first clinical detection on April 4, 2021, by more than 9 months. However, we found first detection of Delta in the wastewater surveillance on April 14, 2021, two and a half months before it was detected in clinical surveillance in Clarke County. Wastewater surveillance also detected Omicron BA.1 on September 1, 2021 and BA.2 on December 6, 2021, three months before they were detected in clinical sequences. The greatest discrepancy was noted in BA.5, which was detected in wastewater on August 18, 2021, about 9 months prior to clinical sequence detection (Supplementary Table 1). These results emphasize a critical advantage of wastewater surveillance in the early detection of VOCs prior to clinical data.

### Variant shifts

TMRCAs for Alpha estimated from wastewater sequences (May 24, 2020, 95% Highest Posterior Density (HPD): January 18 – October 4, 2020), clinical sequences (May 26, 2020, 95% HPD: March 6 – August 17, 2020), and combined data (May 23, 2020, 95% HPD: March 10 – September 2020) were very congruent (Supplementary Table 2, Figure 3). TMRCAs for Delta estimated from wastewater sequences (January 30, 2020, 95% HPD: September 9, 2019 – May 31, 2020), clinical sequences (March 16, 2020, 95% HPD: January 16, 2020 – May 23, 2020), and combined data (February 6, 2020, 95% HPD: December 30, 2019 – March 23, 2020) were also highly congruent. Notably, Delta was inferred to have emerged a few months before the Alpha variant, though there is some overlap in the confidence intervals. Generally, TMRCAs inferred from wastewater sequences had wide confidence intervals for each VOC, likely due to the small sample sizes. While the estimates for the TMRCA were concordant for Alpha and Delta, they were quite different for Omicron sublineages. Specifically, the TMRCA estimates from wastewater sequences were significantly earlier and more uncertain than those estimated from clinical sequences. As expected, the combined analysis provided consistently narrower estimates than the wastewater analysis, and also slightly increased the precision compared to the clinical estimates.

**Figure 3.**
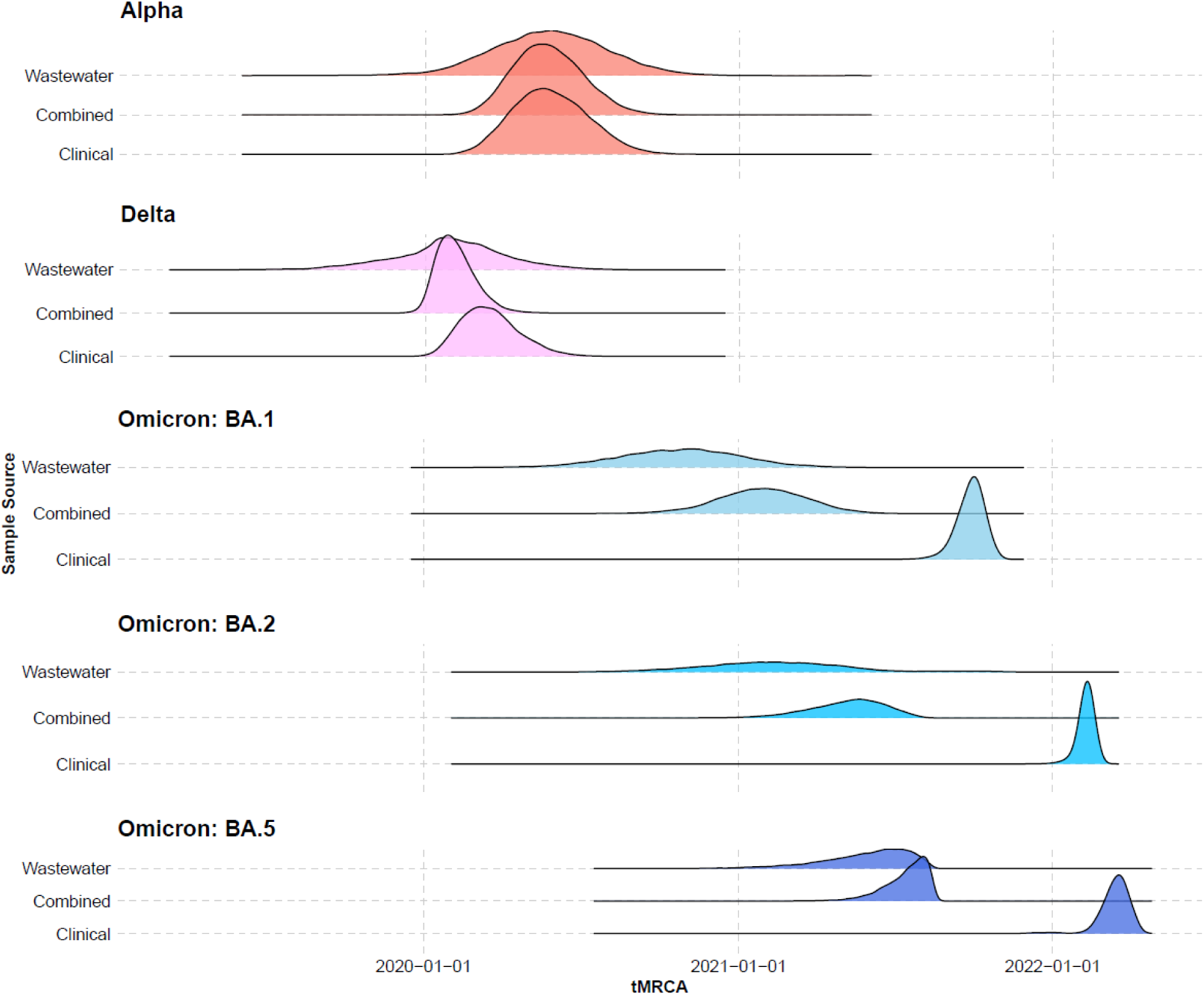
TMRCAs from wastewater sequences are congruent or earlier than clinical sequences. Density plots of the inferred TMRCAs from the log files of the BEAST analyses for wastewater sequences (n = 80), clinical sequences (n = 632), and a combined dataset of clinical and wastewater sequences (n = 712).

Our demographic reconstruction of wastewater data revealed concordance in the overall trajectory, yet exhibited wide uncertainty in effective population size estimates and did not capture all the peaks in population size that were detected from the clinical data (Figure 4). We found that combining data helped reduce uncertainty near the root but smoothed the smaller oscillations in population size captured by clinical genomes, which may contain more mutations and offer more phylogenetic resolution as a result of higher genome coverage.

**Figure 4.**
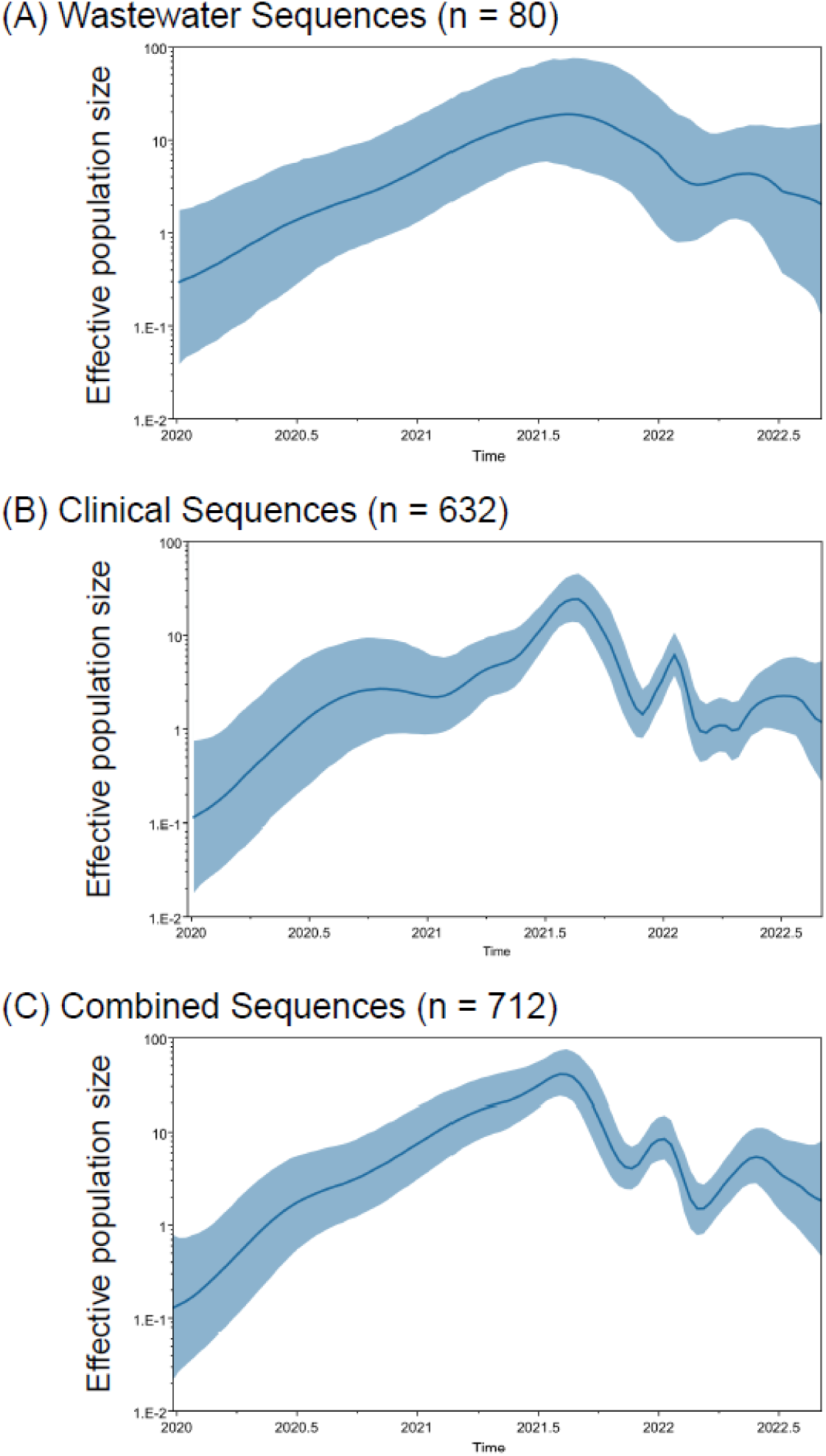
Skyride demographic analyses of wastewater sequences are concordant, yet uncertain. Effective population size (N_e_) over time estimated from ( A) wastewater sequences, (B) clinical sequences, and (C) a combined dataset of clinical and wastewater sequences. Thick solid lines show the mean estimate of effective population size. Light blue shading shows the 95% HPD, with wide intervals demonstrating greater uncertainty.

### Spatial transmission models to study the molecular evolution and underlying mechanisms of viral spread between locations, with the context of important student movement dates

To contextualize the epidemic in Clarke County, we performed a discrete trait analysis. We anticipated that spatial diffusion patterns aligned with longer holiday and university breaks, such as winter, spring, and summer breaks. As students return home or travel, this population movement likely increases connectivity between Clarke County and other locations. It appeared that each clade expansion in the phylogenetic tree began with a university break (Figure 5). This is also reflected in the high number of transitions (Markov jumps) from Clarke County to Georgia and, to a lesser extent, out of state, during time periods in the summer, winter, and spring breaks of 2021 (Supplementary Figure 2). In corroboration, spikes in case counts also appear to be preceded by or occur during a university break (Supplementary Figure 3).

**Figure 5.**
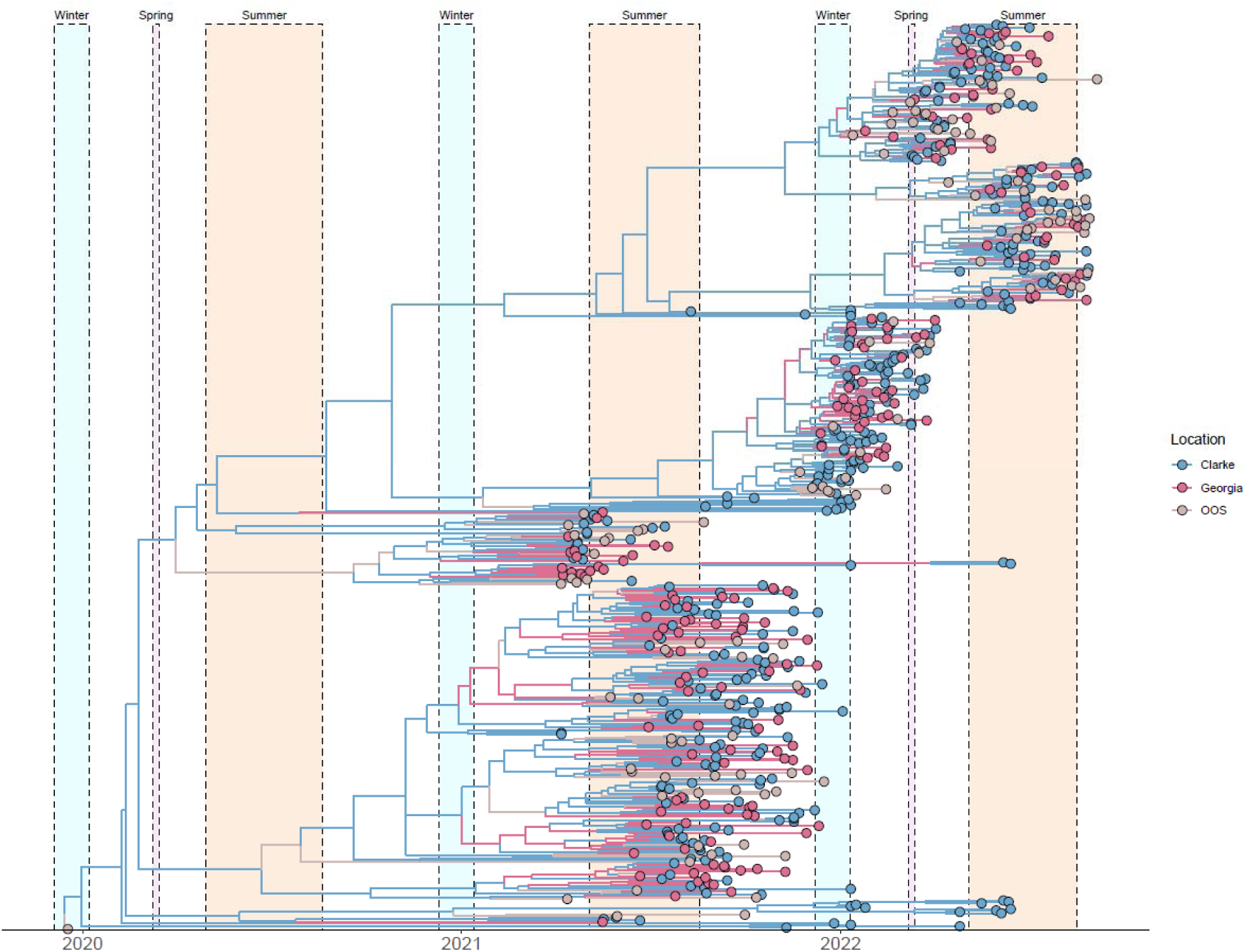
Clade expansion in the context of global transmission and student movement events. Sequences were collected during April 14, 2021 to September 5, 2022 (n=632). Tips and branches are colored by inferred location, including Clarke County (n=341), Georgia (n=182), and out of state (OOS) (n=109). Clade expansion is marked by rapid branching events. Dates of student movement are highlighted, including winter (blue), spring (orange), and summer (pink) break. Spring break in 2021 was canceled due to the COVID-19 pandemic.

We found evidence of a few introductions into Clarke County that led to sustained local transmission. However, the phylogenetic tree revealed substantial mixing between locations, with no distinct spatial structure (Figure 5). The absence of location-specific clades suggests frequent transmission and gene flow between populations. This pattern is consistent with the wastewater-derived sequences, which are dispersed throughout the tree rather than clustering exclusively with clinical sequences from Clarke County (Supplementary Figure 4). Smaller time scales reveal more distinct spatial structuring. Of the 6 possible BSSVS transition rates among locations, we inferred 4 statistically supported (Posterior Probability (PP) > 50%) rates, each with decisive support (BF > 100): Clarke to Georgia at 2.25 transitions/year (PP = 100%), Clarke to out of state at 1.6 transitions/year (PP = 100%), Georgia to Clarke at 1.3 transitions/year (PP = 100%), and out of state to Georgia at 0.48 transitions/year (PP = 99.5%). As a proportion of time represented by the tree, Markov rewards estimated that viruses circulated the longest in Clarke County (67%), followed by Georgia (20%) and out of state (19%). We identified 119 transitions from Clarke County to Georgia compared to 33 transitions from Georgia to Clarke County, as well as 80 transitions from Clarke County to out of state and 2 from out of state to Clarke County (Supplementary Figure 5). These results are consistent with the replicates (Supplemental Figures 6-9).

## Discussion

This study is a result of a multi-year, collaborative project between a multi-disciplinary team across the University of Georgia. All experimental conditions build on existing work, creating methods that will be more efficient to sustainably implement in a variety of settings, including public health and resource-limited laboratories. In this study, we use an optimized platform for SARS-CoV-2 enrichment and sequencing (27) to conduct the first published Bayesian phylogenetic analyses of wastewater sequences. Here, we have demonstrated that wastewater surveillance estimated TMRCAs for Alpha and Delta in Clarke County are congruent with clinical estimates, inferred earlier emergence of Omicron sublineages compared to clinical surveillance, captured all variants from clinical sequencing, detected cryptic Beta transmission not captured by clinical sequences, and identified variants months before detection in clinical sequences. In addition, we have highlighted the shortcomings of wastewater sequences in our study to reconstruct viral demographic history compared to clinical sequences, in terms of uncertainty around population size estimates and identification of viral population peaks/shifts. Lastly, combining wastewater and clinical sequences, we have contextualized spatial diffusion patterns of Clarke County in terms of national and international transmission, and highlighted the potential importance of student movement events in clade expansion, increased transmission, and case surges.

As COVID-19 is removed from the list of reportable diseases, clinical surveillance becomes increasingly unreliable. Wastewater surveillance can fill that gap by providing continuous monitoring when diagnostic testing declines. Wastewater surveillance can be particularly advantageous after waning of random and widescale testing of individuals, or when clinical testing capacity is overwhelmed early in a pandemic or during epidemic surges. While substantial delays can occur between clinical isolate collection, sequencing, and availability of results to public health, wastewater epidemiology provides early indication of COVID-19 surges in the community (15,50). This early warning system for new cases in targeted areas (prior to case reports) provides invaluable time for public health and hospital planning. A key question has been whether variant proportions estimated from wastewater are representative of the sampled population. Interpretation of wastewater surveillance data is potentially confounded in communities with high population movement, including tourists and commuters. However, high congruence has been reported between clinical and wastewater sequences in terms of estimated variant prevalence, circulation patterns, and fitness advantage, suggesting that wastewater-derived estimates are representative of a specific local population (51), even in locations with high levels of commuting (19). Additionally, studies have shown that wastewater variants are more similar to local clinical genotypes than to those from other regions within the US or globally. Wastewater sequences can also identify SARS-CoV-2 genotypes at abundances known to be present in communities. Taken together, there is strong evidence that wastewater samples capture true viral genomic diversity in the sampled population (16).

Phylogenetic analyses of sequences from wastewater have identified novel variants, cryptic community transmission, and identified VOCs up to two weeks before their first detection through clinical surveillance (18,19). Notably, a longitudinal study in India detected Omicron sublineages in wastewater up to two months before the first detection in clinical samples (21). We found that wastewater genomic surveillance identified the three major VOCs (Alpha, Delta, and Omicron) throughout their period of occurrence, even with a small sample size. Wastewater surveillance first detected Delta on April 14, 2021, two and a half months before it was detected in clinical surveillance in Clarke County. Wastewater surveillance also detected Omicron BA.5 on August 18, 2021, four months before any Omicron sublineage was detected in clinical surveillance. Here, wastewater detected Beta, suggesting that it captured cryptic transmission not identified through clinical surveillance alone.

While we found that wastewater is a useful tool to identify and track variants, there are more salient limitations in its use to reconstruct epidemic dynamics and demographic history. Importantly, even with a small sample size, TMRCAs for Alpha and Delta were highly congruent between wastewater and clinical samples. TMRCAs for Omicron sublineages demonstrate that wastewater surveillance is able to detect the emergence of BA.1, BA.2, and BA.5 significantly earlier compared to clinical sequencing, yet the confidence intervals are very wide, likely due to a small sample size for each sublineage. Combining wastewater and clinical sequences consistently narrows confidence intervals compared to individual analyses. When comparing the estimates of population dynamics from wastewater and clinical samples, we found that the skyride plots are aligned in their trajectories, yet the viral effective population sizes (N_e_) inferred from the wastewater sequences exhibit considerable uncertainty and do not capture all of the potential shifts in population size over time. This is possibly due to the inclusion of many low-coverage wastewater sequences that may not contain as much genetic information compared to clinical genomes. Overall, wastewater-derived inferences appear to be largely congruent with clinical sequences, but are less precise. Combining these two sources of data can lead to inferences with more precision, which is necessary to infer parameters in epidemiological dynamics, such the reproductive number, how well control measures are working, or epidemic growing/shrinking. Ultimately, it would be sensible to look into this further with a larger sample size and/or higher coverage wastewater sequences. Comparative analysis of the genomic diversity in wastewater and clinical sequences (e.g., lineage abundance) could help confirm whether wastewater-based inferences accurately reflect local community transmission patterns (e.g., lineage prevalence).

To study viral spread, we applied spatial transmission models to a combined dataset of clinical and wastewater sequences, within the context of national and international transmission and key student movement dates for the University of Georgia. Our results suggest that population movement during university breaks is correlated with clade expansion in the phylogenetic tree, spikes in the number of reported cases, and a higher frequency of transmission from Clarke County to surrounding counties in Georgia. These findings are consistent with mathematical and computational modeling studies showing that student travel impacts transmission dynamics on college campuses, and alternative break schedules can reduce infections (52). Markov jumps and rewards are sensitive to sample sizes; although the dataset was comprised of about a 1:1 ratio of focal to contextual sequences, overrepresentation of Clarke County sequences in relation to Georgia and worldwide sequences likely biased inferences about important sources of transmission towards Clarke County. Future molecular epidemiology studies should also quantitatively assess the impact of student movement on SARS-CoV-2 transmission. For example, McLaughlin et al. (2022) estimated the importation rate of sublineages before and after travel restrictions in Canada to evaluate their effectiveness (53). Similarly, Dellicour et al. (2021) used continuous phylogeographic analysis to assess whether Belgium’s national lockdown altered lineage dispersal velocity (54). Applying such methods to student migration patterns in Clarke County could provide a clearer picture of how mobility-driven introductions contribute to local outbreaks and inform strategies for mitigating future epidemics.

Bioinformatics analysis of high-throughput sequencing data from wastewater is challenging because environmental samples often contain low viral loads, PCR inhibitors, and fragmented RNA, which can lead to poor and inconsistent sequencing coverage and the recovery of only partial genomes (19). In this study, the library quality was likely hindered by the homogenization process, producing very fragmented samples. There is evidence to suggest that vigorous homogenization during RNA extraction may be beneficial for quantification purposes, but detrimental to sequencing outcomes like genomic depth of coverage (55). As a result of the time to optimize our sequencing strategy, our samples degraded in the freezer, leading to small fragments. We lost a third of reads after trimming, another third could not be assigned a strain by Kallisto, and some after filtering by spike protein coverage, resulting in a relatively small sample size of whole genome sequences for downstream phylogenetic analyses. While wastewater surveillance did pick up the presence of Gamma and Mu variants, which were identified in the clinical samples, they were removed during the quality control steps. Due to cost considerations, we sequenced all the samples at once, using one set of tiled amplicon primers. As primers are updated to become more compatible with the latest variants, we lose compatibility with earlier variants. Therefore, we identified a set of primers that would be representative of as many variants across our timeline as possible, resulting in a loss of mu and gamma in our final dataset, as well as a disproportionately low number of Alpha sequences that passed quality control filters. This may explain why the first detection of Alpha in wastewater sequences in our dataset is relatively late, about nine months after clinical detection. Ultimately, this emphasizes the significant impact of primer selection on inferences on variant dating from wastewater surveillance.

Obtaining reliable and high-quality viral sequences from sewage is complicated by each sample being composed of a population of different variants. It is difficult to effectively isolate, enrich, and amplify SARS-CoV-2 target RNA from this complex mixture and reliably distinguish unique genotypes (and their relative abundance). Here, we have optimized viral enrichment and used the best approaches for sequencing viral genome mixtures from the complex sewage environment as described previously (27). Compared to other deconvolution methods (e.g., Freyja and COJAC) that were only validated by ARTIC primers and require short-read sequence data as input, Kallisto is unique in that it allows for all input types (short read and long read) and is validated by multiple sequencing approaches (Freed/Midnight, ARTIC V4, and NEB VarSkip). While Kallisto requires full genomes, we take advantage of the versatile StrainSort pipeline which only needs key areas such as the spike protein region. Due to the challenges in sequencing and assembling wastewater genomes, we built an alignment that is more robust to issues in sequencing errors, misassembles, and other factors.

Privacy concerns around wastewater data collection should also be further explored to ensure that wastewater sequencing remains a viable and ethical surveillance system. Systematic reviews on wastewater are complicated by a lack of standardization across studies. Standardization such as the Environmental Microbiology Minimum Information (EMMI) guidelines should be adopted to support high quality, consistent, and transparent study design in environmental microbiology to improve comparability across studies. These would enhance the translation and relevance of research findings and strengthen the weight of evidence for public health decision-making (56)

Timely sharing of viral genome sequences with key metadata is critical in epidemic surveillance of circulating and emerging variants and inference of SARS-CoV-2 variant dynamics. These phylogenetic analyses provide insight on SARS-CoV-2 circulating variants, revealing variant emergence, population dynamics, and cryptic transmission. Leveraging the strengths of wastewater surveillance can offer a powerful approach for molecular epidemiology that complements the limitations and biases of case-based surveillance. Further guidance on when to focus limited resources on clinical surveillance, wastewater surveillance, or a combined approach are needed.

## Declarations

## Ethics approval and consent to participate

This research has been determined to be exempt by the Human Subjects Office (IRB ID: PROJECT00007534).

## Consent for publication

Not applicable

## Availability of data and materials

All raw wastewater-derived data generated in this study are available under the BioProject accession PRJNA1126268. The protocol for sequencing SARS-CoV-2 in raw wastewater samples is available at https://dx.doi.org/10.17504/protocols.io.rm7vzj6zrlx1/v1. The StrainSort pipeline is available at: https://github.com/mandysulli/StrainSort_pipeline. The dashboard for wastewater surveillance of SARS-CoV-2 in Clarke County is available at https://www.covid19.uga.edu/wastewater-athens.html. Clinical genome sequences and associated metadata are published in GISAID’s EpiCov database, available at https://doi.org/10.55876/gis8.251127nh, https://doi.org/10.55876/gis8.251202nx, https://doi.org/10.55876/gis8.251202hc, https://doi.org/10.55876/gis8.251202st, and Supplementary Data. R scripts are available at https://github.com/Gabriella-Veytsel/wastewater.

## Authors’ contributions

G.V. and J.B conceptualized the study. L.L. and M.L collected wastewater samples and performed library preparation. M.S. performed sequencing of wastewater samples. M.S and G.V. performed genome assembly of wastewater samples. G.V. conducted data analysis, data interpretation, data visualization, and manuscript writing. H.D. contributed resources. J.B., E.L., L.C., and T.G. provided supervision and acquired financial support. All authors contributed to reviewing and editing.

## Acknowledgments

We thank the Georgia Department of Public Health epidemiologists, laboratorians and bioinformaticians. We appreciate the Athens-Clarke County Public Utilities Department for coordinating the collection of all wastewater samples used in this study. We gratefully acknowledge all data contributors, i.e., the Authors and their Originating laboratories responsible for obtaining the specimens, and their Submitting laboratories for generating the genetic sequence and metadata and sharing via the GISAID Initiative, on which this research is based.

---

**Supplementary Figure 1.**
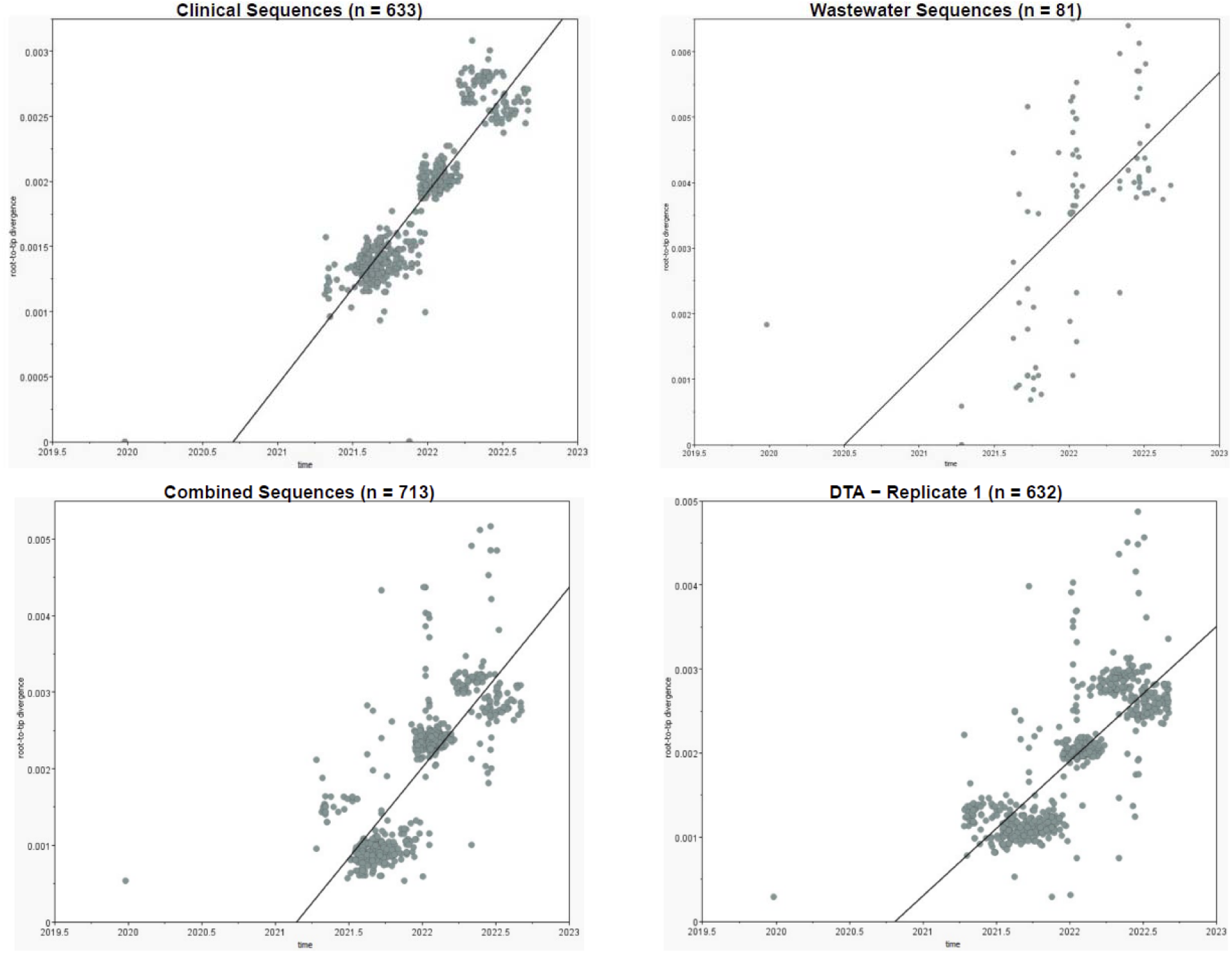
TempEst Root-To-Tip Regression Plots of Clinical and Wastewater Sequences. Divergence from the root of the tree against time of sampling to assess temporal signal in data.

**Supplementary Figure 2.**
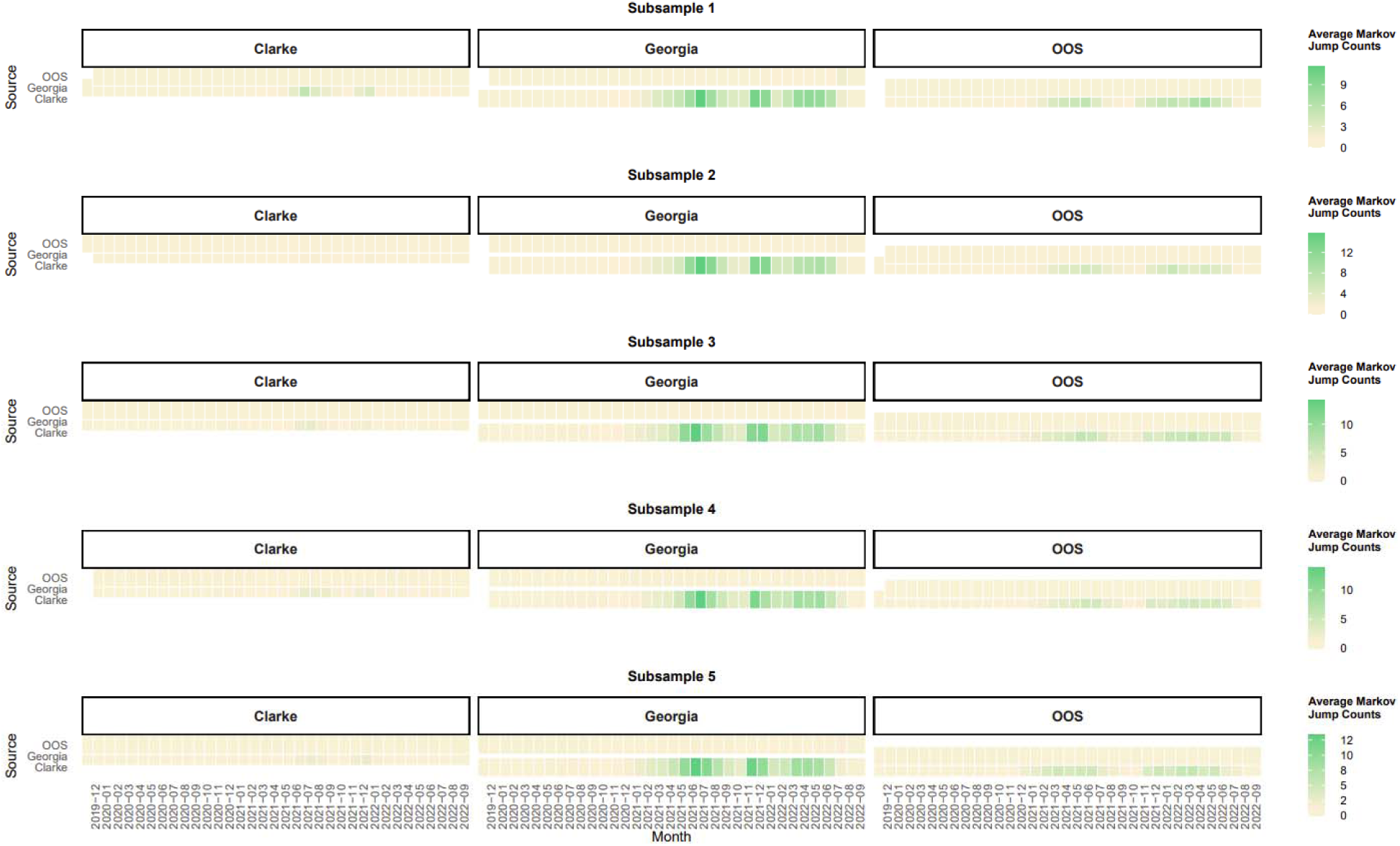
Transitions between locations over time. Phylogenetic discrete trait analyses inferred transitions (Markov jumps) between locations over time along phylogenetic branches across five replicates. The majority of transitions occurred from Clarke County to Georgia and, to a lesser extent, Clarke County to out of state (OOS). These results are consistent within replicates.

**Supplementary Figure 3.**
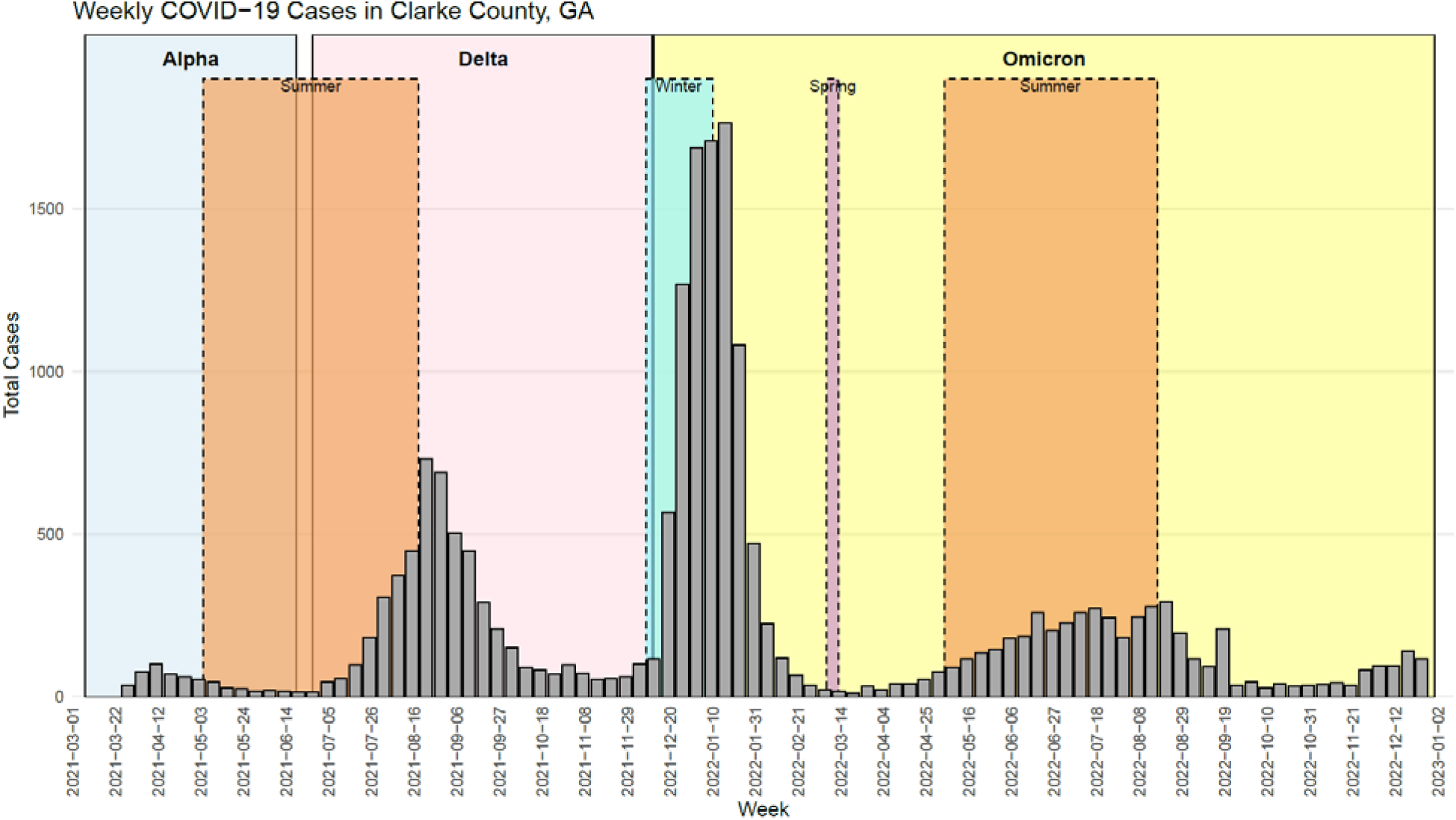
COVID-19 case counts in Clarke County, Georgia. Case surges in the context of key student movement dates and epidemic waves (Alpha, Delta, and Omicron). The dates of variant waves were estimated from clinical genomes in Georgia based on the time periods of dominance compared to other circulating lineages. The Alpha wave lasted between March 7, 2021 – June 19, 2021, the Delta wave lasted between June 27, 2021 – December 11, 2021, and the Omicron wave began on December 12, 2021, and continued past the study period.

**Supplementary Figure 4.**
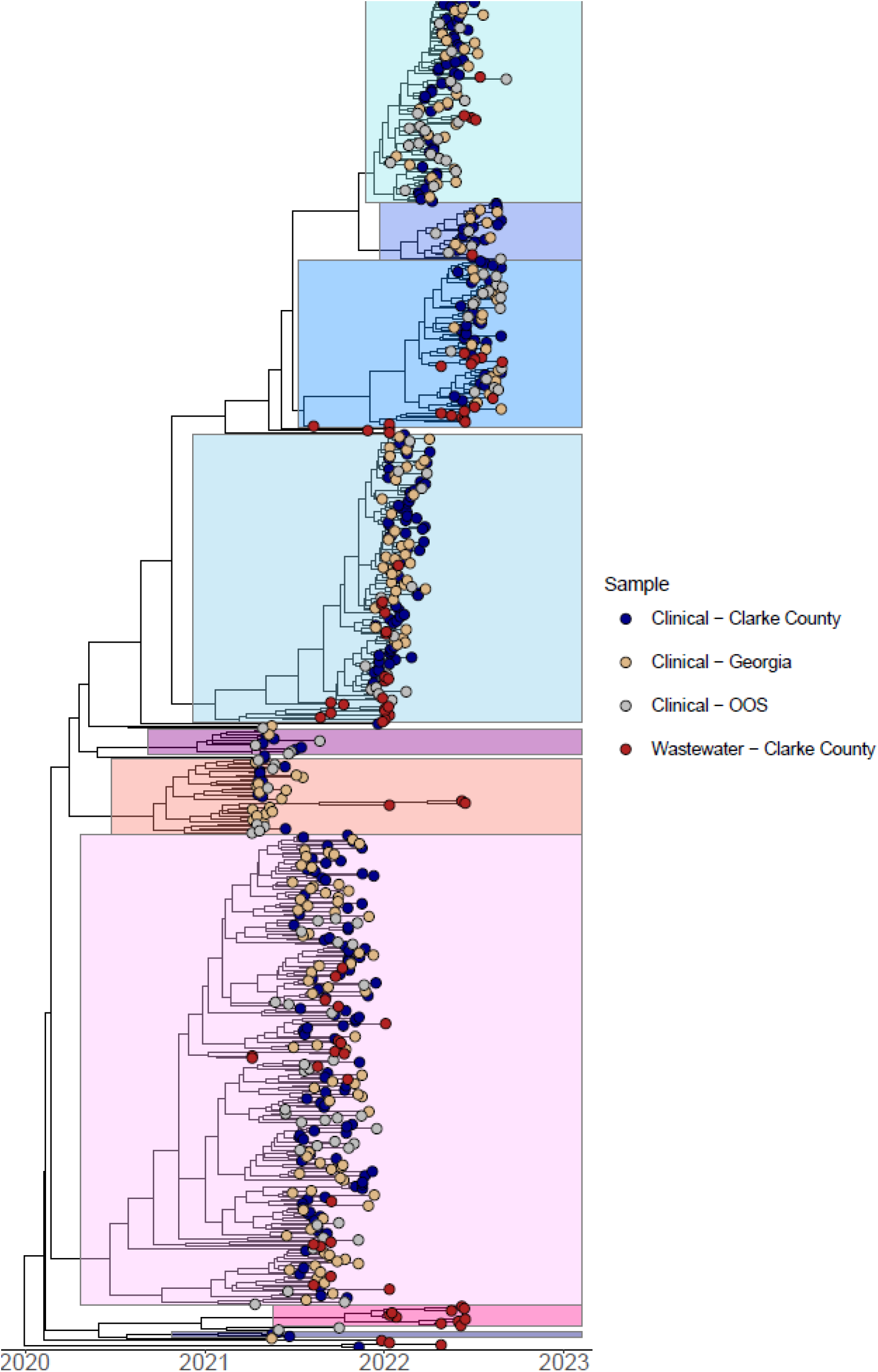
Combined clinical and wastewater phylogenetic tree, global. Bayesian phylogenetic discrete trait analysis of whole genome sequences collected from clinical and wastewater samples in Clarke County, Georgia with contextual sequences (n = 632). This adjusts Figure 5 by coloring the tips by sample source and highlighting variants to examine the clustering of wastewater sequences with clinical sequences from various locations. Wastewater sequences are dispersed throughout the tree rather than clustering exclusively with clinical sequences from Clarke County.

**Supplementary Figure 5.**
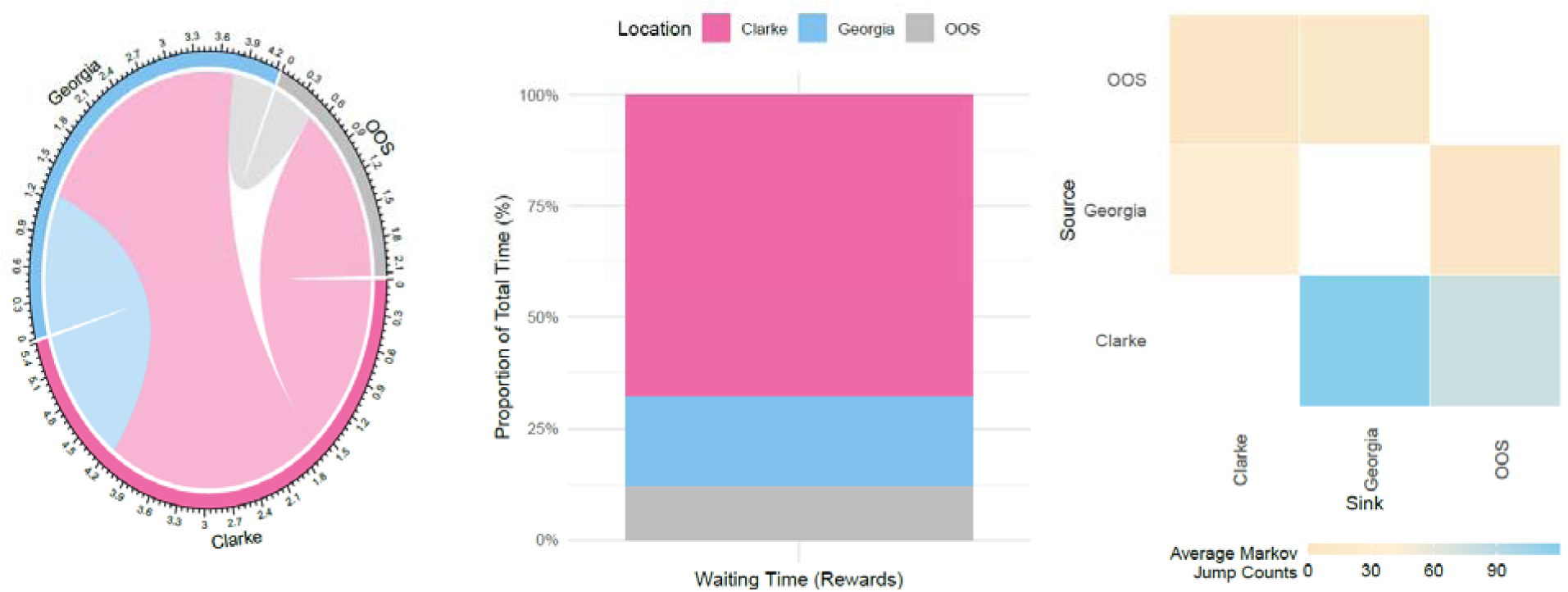
Phylogenetic discrete trait analyses. Inferred BSSVS transition rates (transitions per year), the waiting time (Markov rewards) spent in each location between transitions, and transitions (Markov jumps) between locations along phylogenetic branches. (A) The magnitude of the viral transition rate is proportional to the width of the band. Rates were inferred using BSSVS. Out of the 6 possible transitions, we inferred 4 statistically supported (Posterior Probability (PP) > 50%), each with decisive support (BF > 100): Clarke to Georgia, Clarke to out of state, Georgia to Clarke, and out of state to Georgia. (B) Viruses circulated in Clarke County the majority of the time. (C) The vast majority of transitions occurred from Clarke County to Georgia and, to a lesser extent, Clarke County to out of state (OOS). These results are consistent with replicates.

**Supplementary Figure 6.**
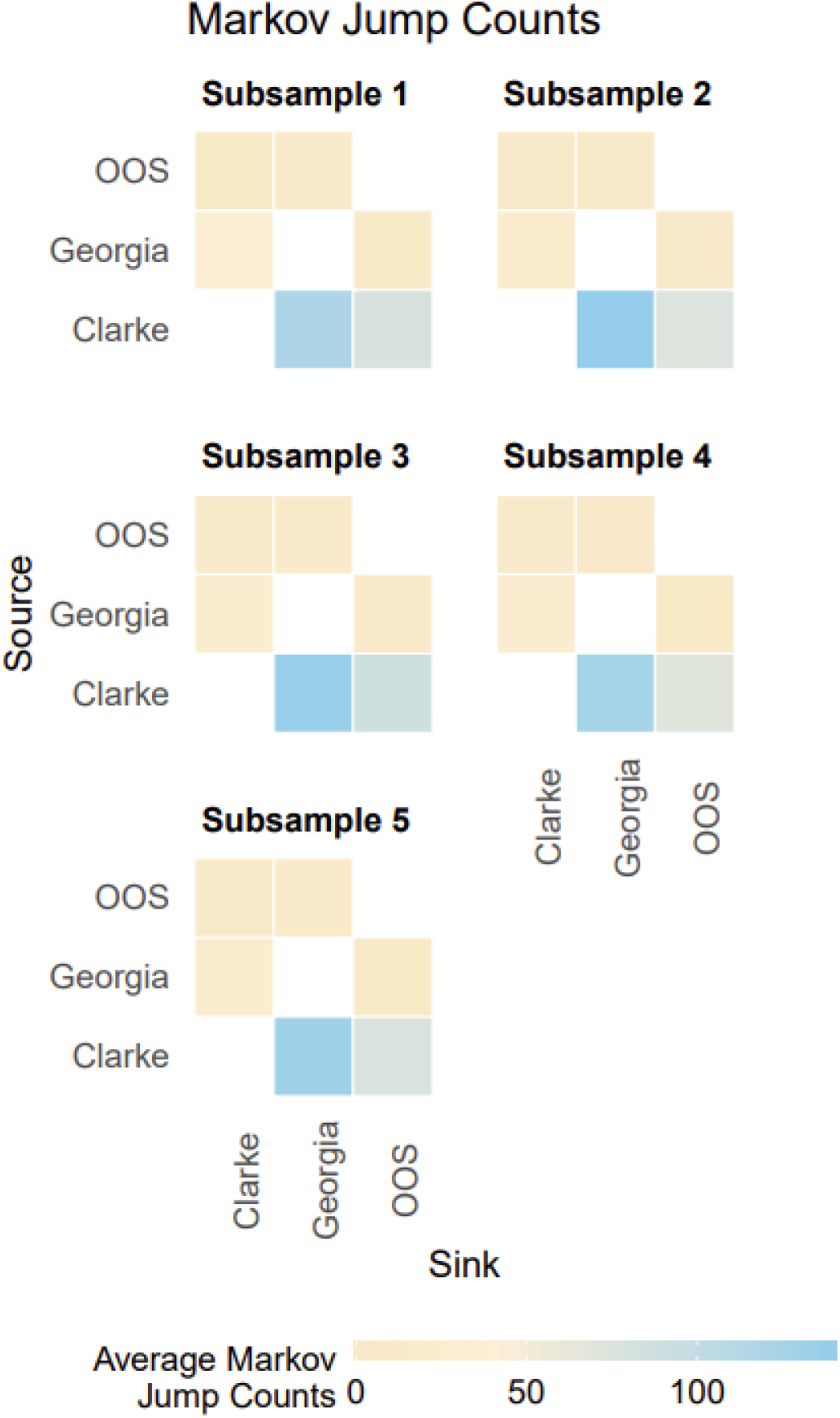
Average Markov jump counts across replicates. Phylogenetic discrete trait analyses inferred transitions (Markov jumps) between locations along phylogenetic branches across five replicates. The majority of transitions occurred from Clarke County to Georgia and, to a lesser extent, Clarke County to out of state (OOS). These results are consistent within replicates.

**Supplementary Figure 7.**
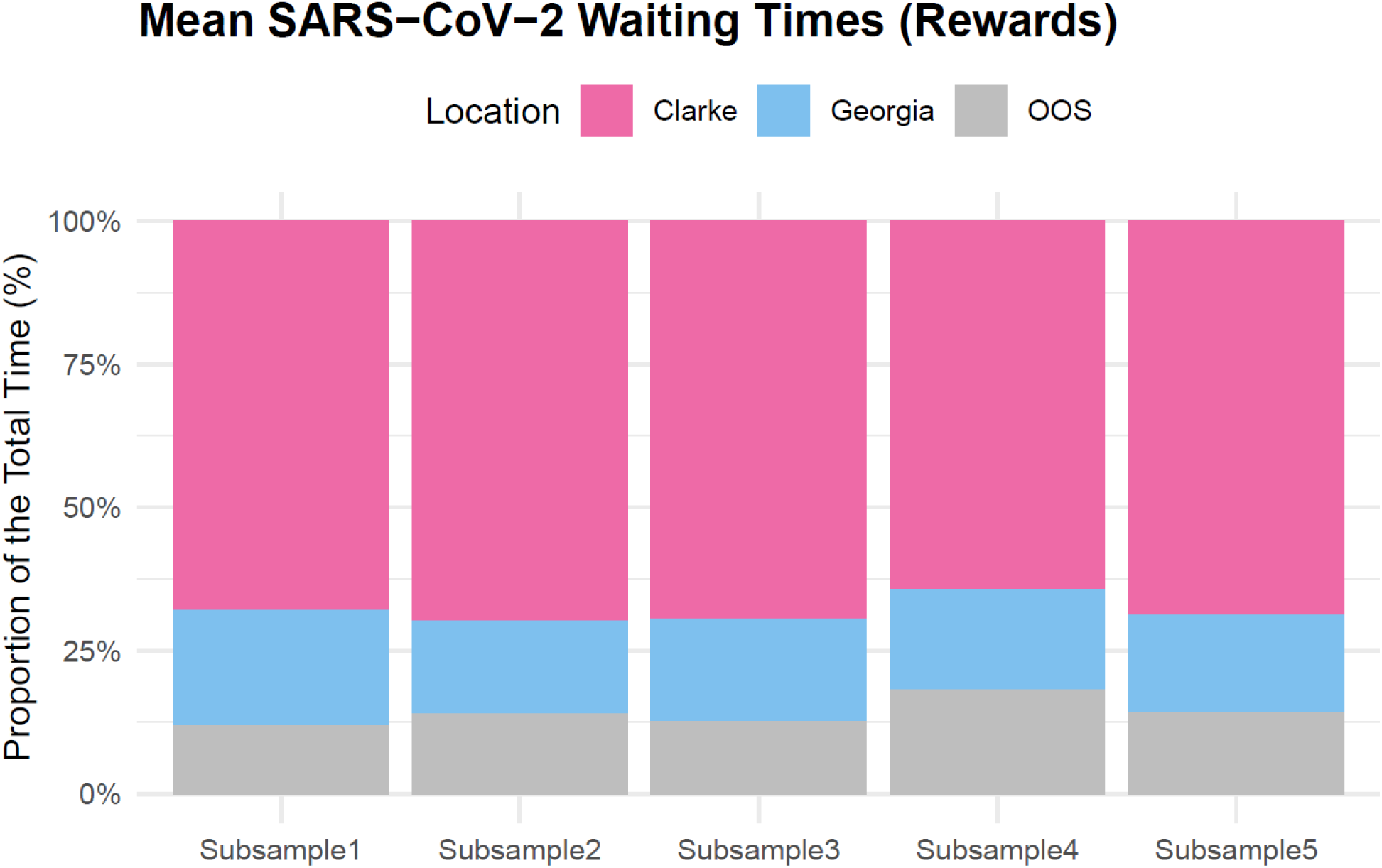
Mean Markov rewards across replicates. Phylogenetic discrete trait analyses inferred the waiting time (Markov rewards) spent in each location between transitions across five replicates. Consistent across replicates, viruses circulated in Clarke County the majority of the time.

**Supplementary Figure 8.**
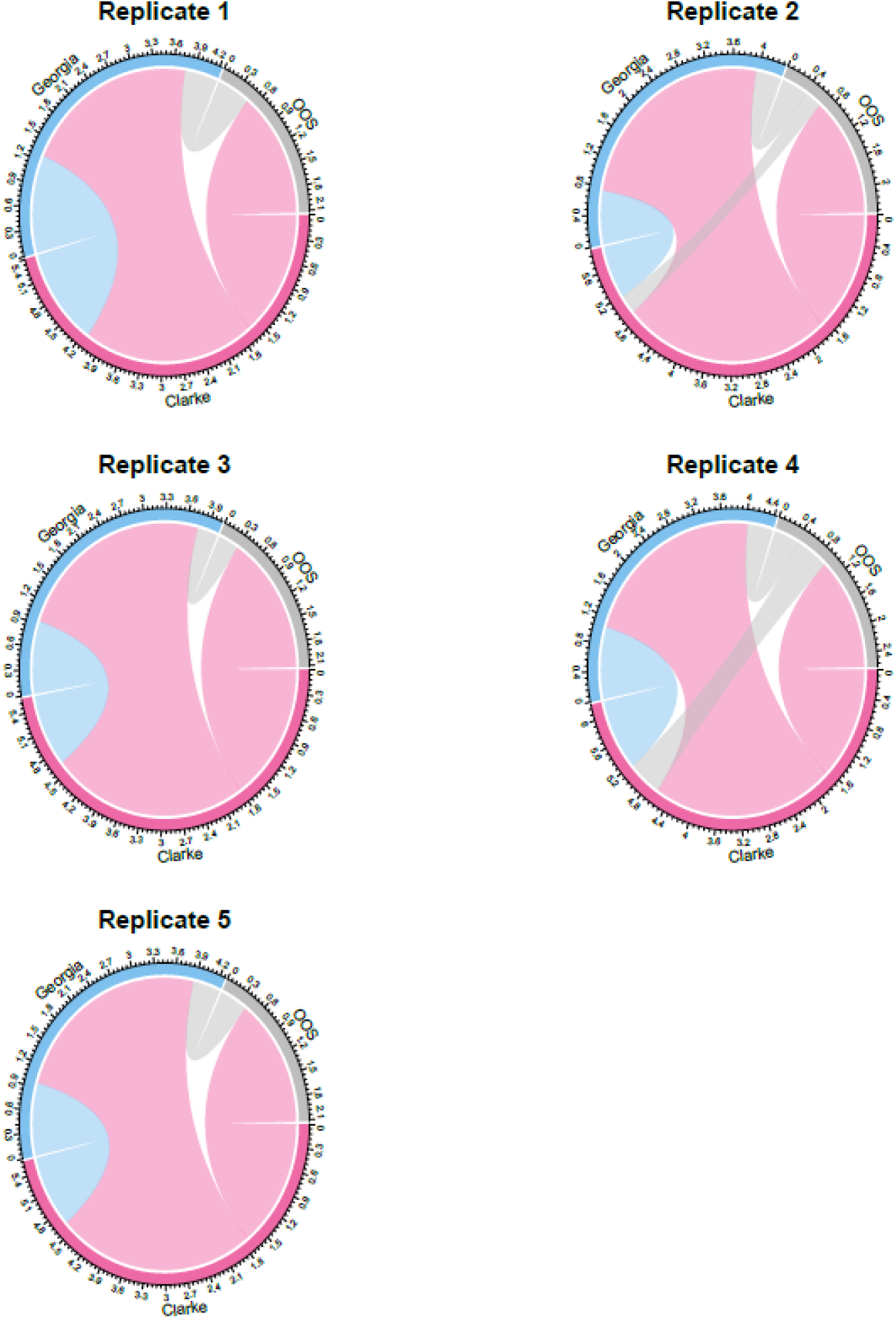
BSSVS transition rates across replicates. Phylogenetic discrete trai analyses inferred BSSVS transition rates (transitions per year) across five replicates. Out of the 6 possible transitions, we inferred 4 statistically supported (Posterior Probability (PP) > 50%), each with decisive support (BF > 100): Clarke to Georgia, Clarke to out of state, Georgia to Clarke, and out of state to Georgia. The transition rate from out of state to Clarke was found to be statistically supported for two replicates, but only substantial support was estimated (BF = 3-10).

**Supplementary Figure 9.**
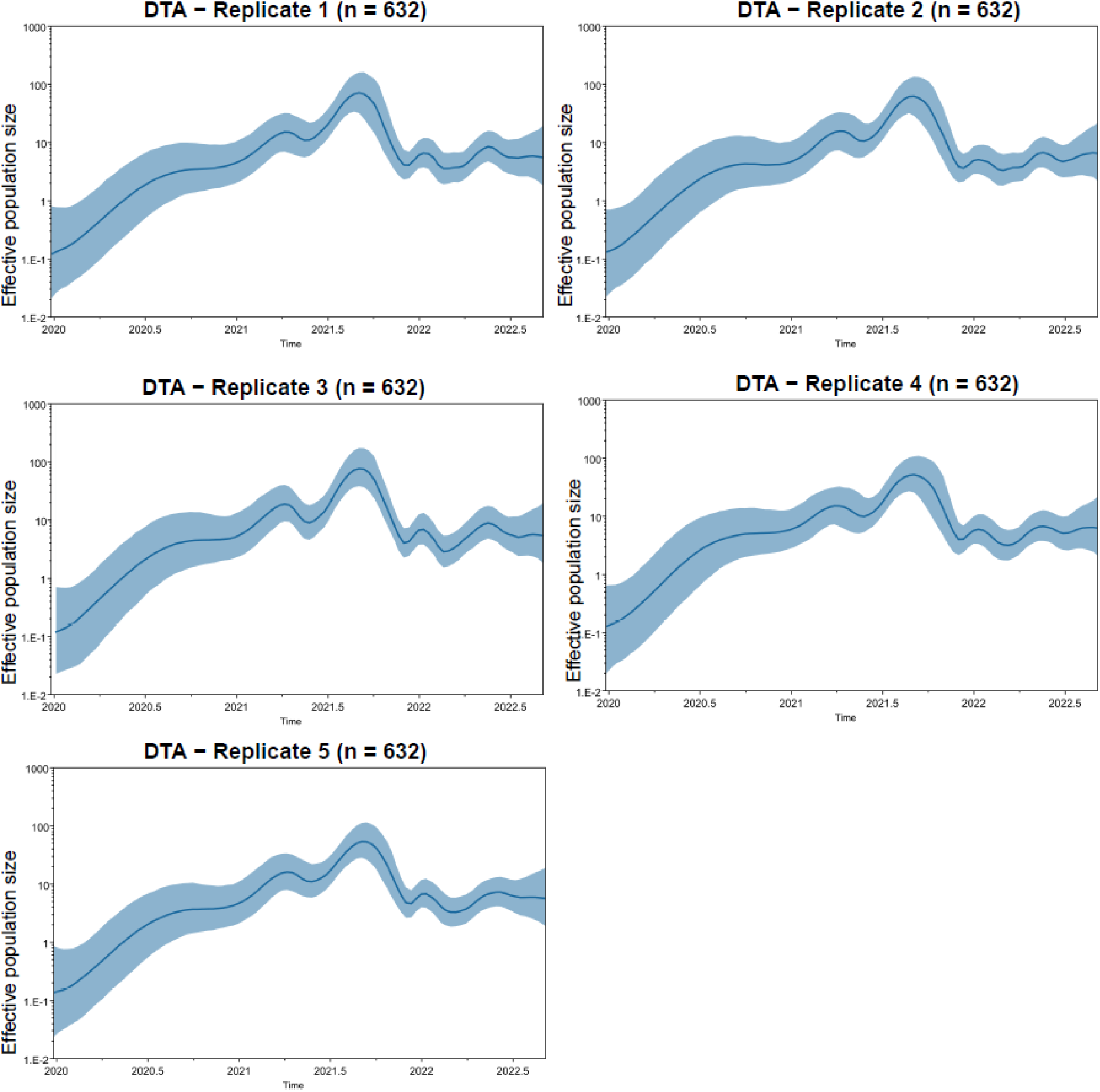
Demographic analyses across replicates. Effective population size (N_e_) over time estimated from a combined dataset of focal and contextual sequences from Georgia and worldwide.

## References

1. Brito AF, Semenova E, Dudas G, Hassler GW, Kalinich CC, Kraemer MUG, et al. Global disparities in SARS-CoV-2 genomic surveillance. Nat Commun. 2022 Nov 16;13(1):7003.

2. World Health Organization. Genomic sequencing of SARS-CoV-2: a guide to implementation for maximum impact on public health [Internet]. Genève: World Health Organization; 2022 [cited 2022 Nov 1]. Available from: https://apps.who.int/iris/handle/10665/360580

3. Li L, Uppal T, Hartley PD, Gorzalski A, Pandori M, Picker MA, et al. Detecting SARS-CoV-2 variants in wastewater and their correlation with circulating variants in the communities. Scientific Reports. 2022 Sept 27;12:16141.

4. Michie A. Wastewater-based SARS-CoV-2 surveillance and sequencing. Microbiol Aust. 2024 Mar 13;45(1):8–12.

5. Melnick JL. POLIOMYELITIS VIRUS IN URBAN SEWAGE IN EPIDEMIC AND IN NONEPIDEMIC TIMES1. American Journal of Epidemiology. 1947 Mar;45(2):240–53.

6. Mercier E, D’Aoust PM, Thakali O, Hegazy N, Jia JJ, Zhang Z, et al. Municipal and neighbourhood level wastewater surveillance and subtyping of an influenza virus outbreak. Sci Rep. 2022 Sept 22;12(1):15777.

7. Ando H, Ahmed W, Iwamoto R, Ando Y, Okabe S, Kitajima M. Impact of the COVID-19 pandemic on the prevalence of influenza A and respiratory syncytial viruses elucidated by wastewater-based epidemiology. Science of The Total Environment. 2023 July 1;880:162694.

8. Bar-Or I, Indenbaum V, Weil M, Elul M, Levi N, Aguvaev I, et al. National Scale Real-Time Surveillance of SARS-CoV-2 Variants Dynamics by Wastewater Monitoring in Israel. Viruses. 2022 June;14(6):1229.

9. Tiwari A, Adhikari S, Kaya D, Islam MdA, Malla B, Sherchan SP, et al. Monkeypox outbreak: Wastewater and environmental surveillance perspective. Science of The Total Environment. 2023 Jan 15;856:159166.

10. Wu F, Oghuan J, Gitter A, Mena KD, Brown EL. Wide mismatches in the sequences of primers and probes for monkeypox virus diagnostic assays. Journal of Medical Virology. 2023;95(1):e28395.

11. CDC. Centers for Disease Control and Prevention. 2023 [cited 2025 Feb 10]. National Wastewater Surveillance System. Available from: https://www.cdc.gov/nwss/wastewater-surveillance.html

12. Deslandes A, Berti V, Tandjaoui-Lambotte Y, Alloui C, Carbonnelle E, Zahar JR, et al. SARS-CoV-2 was already spreading in France in late December 2019. International Journal of Antimicrobial Agents. 2020 June 1;55(6):106006.

13. La Rosa G, Mancini P, Bonanno Ferraro G, Veneri C, Iaconelli M, Bonadonna L, et al. SARS-CoV-2 has been circulating in northern Italy since December 2019: Evidence from environmental monitoring. Science of The Total Environment. 2021 Jan 1;750:141711.

14. Randazzo W, Truchado P, Cuevas-Ferrando E, Simón P, Allende A, Sánchez G. SARS-CoV-2 RNA in wastewater anticipated COVID-19 occurrence in a low prevalence area. Water Res. 2020 Aug 15;181:115942.

15. Weidhaas J, Aanderud ZT, Roper DK, VanDerslice J, Gaddis EB, Ostermiller J, et al. Correlation of SARS-CoV-2 RNA in wastewater with COVID-19 disease burden in sewersheds. Science of The Total Environment. 2021 June 25;775:145790.

16. Crits-Christoph A, Kantor RS, Olm MR, Whitney ON, Al-Shayeb B, Lou YC, et al. Genome Sequencing of Sewage Detects Regionally Prevalent SARS-CoV-2 Variants. mBio. 2021 Jan 19;12(1):10.1128/mbio.02703-20.

17. Izquierdo-Lara R, Elsinga G, Heijnen L, Munnink BBO, Schapendonk CME, Nieuwenhuijse D, et al. Monitoring SARS-CoV-2 Circulation and Diversity through Community Wastewater Sequencing, the Netherlands and Belgium. Emerg Infect Dis. 2021 May;27(5):1405–15.

18. Karthikeyan S, Ronquillo N, Belda-Ferre P, Alvarado D, Javidi T, Longhurst CA, et al. High-Throughput Wastewater SARS-CoV-2 Detection Enables Forecasting of Community Infection Dynamics in San Diego County. mSystems. 2021 Mar 2;6(2):e00045–21.

19. Jahn K, Dreifuss D, Topolsky I, Kull A, Ganesanandamoorthy P, Fernandez-Cassi X, et al. Early detection and surveillance of SARS-CoV-2 genomic variants in wastewater using COJAC. Nat Microbiol. 2022 Aug;7(8):1151–60.

20. Baaijens JA, Zulli A, Ott IM, Nika I, van der Lugt MJ, Petrone ME, et al. Lineage abundance estimation for SARS-CoV-2 in wastewater using transcriptome quantification techniques. Genome Biology. 2022 Nov 8;23(1):236.

21. Lamba S, Ganesan S, Daroch N, Paul K, Joshi SG, Sreenivas D, et al. SARS-CoV-2 infection dynamics and genomic surveillance to detect variants in wastewater – a longitudinal study in Bengaluru, India. The Lancet Regional Health - Southeast Asia [Internet]. 2023 Apr 1 [cited 2024 Oct 16];11. Available from: https://www.thelancet.com/journals/lansea/article/PIIS2772-3682(23)00011-2/fulltext

22. Wang T, Wang C, Myshkevych Y, Mantilla-Calderon D, Talley E, Hong PY. SARS-CoV-2 wastewater-based epidemiology in an enclosed compound: A 2.5-year survey to identify factors contributing to local community dissemination. Science of The Total Environment. 2023 June 1;875:162466.

23. Férez JA, Cuevas-Ferrando E, Nicolás MAS, Andreu PJS, López R, Truchado P, et al. Wastewater-Based Epidemiology to Describe the Evolution of SARS-CoV-2 in the South-East of Spain, and Application of Phylogenetic Analysis and a Machine Learning Approach. Viruses. 2023 July 3;15(7):1499.

24. Nemudryi A, Nemudraia A, Wiegand T, Surya K, Buyukyoruk M, Cicha C, et al. Temporal Detection and Phylogenetic Assessment of SARS-CoV-2 in Municipal Wastewater. CR Med [Internet]. 2020 Sept 22 [cited 2025 Feb 27];1(6). Available from: https://www.cell.com/cell-reports-medicine/abstract/S2666-3791(20)30124-5

25. Lariscy LM, Lott ME, Handel A, Foley AM, Melendez-Declet C, Metsker L, et al. Balancing Accuracy and Actionability: An Assessment of Minimal-Input Wastewater Models for COVID-19 Prediction [Internet]. medRxiv; 2025 [cited 2025 July 7]. p. 2025.07.03.25330828. Available from: https://www.medrxiv.org/content/10.1101/2025.07.03.25330828v1

26. Lott MEJ, Norfolk WA, Dailey CA, Foley AM, Melendez-Declet C, Robertson MJ, et al. Direct wastewater extraction as a simple and effective method for SARS-CoV-2 surveillance and COVID-19 community-level monitoring. FEMS Microbes. 2023;4:xtad004.

27. Lott MEJ, Sullivan AH, Lariscy LM, Norfolk WA, Dillon KC, Beaudry MS, et al. Comparison of three tiled amplicon sequencing approaches for SARS-CoV-2 variant detection from wastewater [Internet]. 2024 [cited 2025 Feb 25]. Available from: http://biorxiv.org/lookup/doi/10.1101/2024.06.16.599198

28. Goswami K, Sanan-Mishra N. RNA-seq for revealing the function of the transcriptome. In: Bioinformatics [Internet]. Elsevier; 2022 [cited 2025 Feb 26]. p. 105–29. Available from: https://linkinghub.elsevier.com/retrieve/pii/B978032389775400002X

29. Bolger AM, Lohse M, Usadel B. Trimmomatic: a flexible trimmer for Illumina sequence data. Bioinformatics. 2014 Aug 1;30(15):2114–20.

30. Sullivan AH. MINING MIXTURES: APPROACHES TO ENRICH AND DETECT PATHOGEN SEQUENCES IN COMPLEX SAMPLES TO IMPROVE PUBLIC HEALTH SURVEILLANCE [Internet] [Dissertation]. [Athens, GA]: University of Georgia; 2023. Available from: https://openscholar.uga.edu/record/2539#files

31. Prjibelski A, Antipov D, Meleshko D, Lapidus A, Korobeynikov A. Using SPAdes De Novo Assembler. CP in Bioinformatics. 2020 June;70(1):e102.

32. Shu Y, McCauley J. GISAID: Global initiative on sharing all influenza data – from vision to reality. Eurosurveillance. 2017 Mar 30;22(13):30494.

33. Katoh K, Standley DM. MAFFT Multiple Sequence Alignment Software Version 7: Improvements in Performance and Usability. Molecular Biology and Evolution. 2013 Apr 1;30(4):772–80.

34. Alisoltani A, Jaroszewski L, Iyer M, Iranzadeh A, Godzik A. Increased Frequency of Indels in Hypervariable Regions of SARS-CoV-2 Proteins—A Possible Signature of Adaptive Selection. Front Genet [Internet]. 2022 June 2 [cited 2025 Feb 27];13. Available from: https://www.frontiersin.org/journals/genetics/articles/10.3389/fgene.2022.875406/full

35. Badua CLDC, Baldo KAT, Medina PMB. Genomic and proteomic mutation landscapes of SARS-CoV-2. Journal of Medical Virology. 2021;93(3):1702–21.

36. Chrisman BS, Paskov K, Stockham Nate, Tabatabaei K, Jung JY, Washington P, et al. Indels in SARS-CoV-2 occur at template-switching hotspots. BioData Mining. 2021 Mar 20;14(1):20.

37. Minh BQ, Lanfear R, Ly-Trong N, Trifinopoulos J, Schrempf D, Schmidt HA. IQ-TREE version 2.2.0: Tutorials and Manual Phylogenomic software by maximum likelihood [Internet]. 2022. Available from: http://www.iqtree.org/doc/iqtree-doc.pdf

38. Rambaut A, Lam TT, Max Carvalho L, Pybus OG. Exploring the temporal structure of heterochronous sequences using TempEst (formerly Path-O-Gen). Virus Evol. 2016 Jan;2(1):vew007.

39. Suchard MA, Lemey P, Baele G, Ayres DL, Drummond AJ, Rambaut A. Bayesian phylogenetic and phylodynamic data integration using BEAST 1.10. Virus Evolution [Internet]. 2018 Jan 1 [cited 2022 Nov 2];4(1). Available from: https://academic.oup.com/ve/article/doi/10.1093/ve/vey016/5035211

40. Rambaut A, Holmes EC, O’Toole Á, Hill V, McCrone JT, Ruis C, et al. A dynamic nomenclature proposal for SARS-CoV-2 lineages to assist genomic epidemiology. Nat Microbiol. 2020 Nov;5(11):1403–7.

41. Rambaut A, Drummond AJ, Xie D, Baele G, Suchard MA. Posterior Summarization in Bayesian Phylogenetics Using Tracer 1.7. Susko E, editor. Systematic Biology. 2018 Sept 1;67(5):901–4.

42. Lemey P, Rambaut A, Drummond AJ, Suchard MA. Bayesian phylogeography finds its roots. PLoS Comput Biol. 2009 Sept;5(9):e1000520.

43. Bielejec F, Baele G, Vrancken B, Suchard MA, Rambaut A, Lemey P. SpreaD3: Interactive Visualization of Spatiotemporal History and Trait Evolutionary Processes. Mol Biol Evol. 2016 Aug;33(8):2167–9.

44. Jeffreys H. Theory of Probability. 3rd ed. Oxford Univ. Press: Oxford; 1961. (Oxford Classic Texts in the Physical Sciences).

45. Kass RE, Raftery AE. Bayes Factors. Journal of the American Statistical Association. 1995 June;90(430):773–95.

46. Hill V, Du Plessis L, Peacock TP, Aggarwal D, Colquhoun R, Carabelli AM, et al. The origins and molecular evolution of SARS-CoV-2 lineage B.1.1.7 in the UK. Virus Evolution. 2022 Sept 21;8(2):veac080.

47. McCrone JT, Hill V, Bajaj S, Pena RE, Lambert BC, Inward R, et al. Context-specific emergence and growth of the SARS-CoV-2 Delta variant. Nature. 2022 Oct 6;610(7930):154–60.

48. Viana R, Moyo S, Amoako DG, Tegally H, Scheepers C, Althaus CL, et al. Rapid epidemic expansion of the SARS-CoV-2 Omicron variant in southern Africa. Nature. 2022 Mar;603(7902):679–86.

49. Meredith M, Kruschke J. HDInterval: Highest (Posterior) Density Intervals [Internet]. 2016 [cited 2025 Jan 28]. p. 0.2.4. Available from: https://CRAN.R-project.org/package=HDInterval

50. Bastardo-Méndez M, Rangel HR, Pujol FH, Grillet ME, Jaspe RC, Malaver N, et al. Detection of SARS-CoV-2 in wastewater as an earlier predictor of COVID-19 epidemic peaks in Venezuela. Sci Rep. 2024 Nov 8;14(1):27294.

51. Agrawal S, Orschler L, Schubert S, Zachmann K, Heijnen L, Tavazzi S, et al. Prevalence and circulation patterns of SARS-CoV-2 variants in European sewage mirror clinical data of 54 European cities. Water Research. 2022 May 1;214:118162.

52. Lehnig CL, Oren E, Vaidya NK. Effectiveness of alternative semester break schedules on reducing COVID-19 incidence on college campuses. Sci Rep. 2022 Feb 8;12(1):2116.

53. McLaughlin A, Montoya V, Miller RL, Mordecai GJ, Canadian COVID-19 Genomics Network (CanCOGen) Consortium, Worobey M, et al. Genomic epidemiology of the first two waves of SARS-CoV-2 in Canada. eLife. 2022 Aug 2;11:e73896.

54. Dellicour S, Durkin K, Hong SL, Vanmechelen B, Martí-Carreras J, Gill MS, et al. A Phylodynamic Workflow to Rapidly Gain Insights into the Dispersal History and Dynamics of SARS-CoV-2 Lineages. Molecular Biology and Evolution. 2021 Apr 1;38(4):1608–13.

55. Feng S, Owens SM, Shrestha A, Poretsky R, Hartmann EM, Wells G. Intensity of sample processing methods impacts wastewater SARS-CoV-2 whole genome amplicon sequencing outcomes. Science of The Total Environment. 2023 June 10;876:162572.

56. Borchardt MA, Boehm AB, Salit M, Spencer SK, Wigginton KR, Noble RT. The Environmental Microbiology Minimum Information (EMMI) Guidelines: qPCR and dPCR Quality and Reporting for Environmental Microbiology. Environ Sci Technol. 2021 Aug 3;55(15):10210–23.

